# Polygenicity at the pathway level for anorexia nervosa

**DOI:** 10.1101/2025.10.08.25337623

**Authors:** Jiayi Xu, Jessica S. Johnson, Chaoyu Liu, Judit García-González, Shing Wan Choi, Eating Disorders Working Group of the Psychiatric Genomics Consortium (PGC-ED), Kristen J. Brennand, Cynthia M. Bulik, Paul F. O’Reilly, Laura M. Huckins

## Abstract

Genome-wide association studies of common disorders reveal polygenic architectures. While these variants may converge into biologically relevant pathways, how polygenicity aggregates across pathways remains unknown. We studied polygenicity at the pathway level in anorexia nervosa (AN)—a psychiatric disorder with multi-systemic clinical manifestations. We constructed pathway-based polygenic risk scores (pathway PRS) to model genetic risk at the pathway level for 3,687 individuals with AN and 11,257 controls. We identified 497 AN-associated pathways after Bonferroni correction, including pathways involved in the brain, metabolism, immunity/stress, and reproduction/development. A strong positive correlation was observed between number of pathways ranking at top 10% and AN case proportion (*r* = 0.74, P = 7.98×10^−21^). The positive correlation between pathway count and AN risk was also observed when restricted to individuals with low overall genetic risk of AN. Pathways ranking at top 10% among cases were more diverse than those in controls. Higher AN risk was observed for pathway aggregation within function (e.g., brain or metabolism) and across functions (e.g., brain + metabolism). Individuals at top 10% risk of brain-brain and brain-metabolism pathway pairs had the highest average AN risk compared to other pathway pairs (e.g., brain-immune, metabolism- metabolism). Further, pathway PRS provided higher prediction power of AN than overall genome-wide PRS and contributed to the genetic liability of AN among AN cases with low overall genetic risk. Together, our results demonstrate that polygenicity exists at the pathway level, opening new avenues for disease prediction and identification of actionable targets.

## Introduction

Understanding the genetic landscape of complex traits requires consideration of polygenicity. The majority of psychiatric traits and disorders have demonstrated polygenic underpinnings^1–6^; meaning that individuals with higher genome-wide burdens of risk alleles are more likely to display the trait. Further, individuals with high numbers of risk alleles for a certain disorder (for example, schizophrenia^7^ or anorexia nervosa^8,9^) are more likely to experience disorder-associated symptoms or characteristics, even in the absence of the disorder itself (for example, psychosis^7^ or weight loss^8,9^). These patterns hold across populations and disorders, and display dose-dependent effects, such that individuals become more likely to exhibit a trait as genetic risk increases^10,11^. Thus, aggregation of genome-wide risk variants clearly has relevance to disease development. However, despite population-level efficacy, polygenic risk scores (PRS) for psychiatric traits and disorders (calculated as a sum of genome-wide risk alleles, weighted by their effect size) have only low predictive power on an individual level^5^; we cannot predict the outcome of any one patient based on PRS. Even among diseases with high heritability^12,13^, many unaffected individuals (controls) have high PRS, while many cases have low PRS, hampering the clinical utility of PRS.

A core issue hindering the application of PRS to the clinic is a lack of understanding of the underlying pathways through which polygenic risk confers disease risk. On a broad scale, we do not understand why large numbers of low-effect-size risk alleles confer disease risk, especially when only a small proportion of them are significantly associated loci. Moreover, we cannot predict, based on PRS, who will develop disease. Major questions remain outstanding. For example, does localization within specific pathways matter more than overall number of variants? If so, does the number or type of pathways matter? And is the effect of genetic risk on disease stronger when variants are concentrated within a few pathways, or spread across the genome?

Our approach applies pathway-based polygenic risk scores (pathway PRS) to genotype data from cases with AN and controls, collected as part of the Psychiatric Genomics Consortium Eating Disorders (PGC-ED) Working Group. AN is an ideal test trait for this approach, for several complementary reasons. First, AN is a multi-systemic trait, with demonstrated metabolic and psychiatric involvement^14^. The role of these systems is borne out in symptomatology^15^, diagnostic criteria^16^, comorbidities^17,18^, and our own previous genetic studies^19,20^. From a metabolic perspective, individuals with AN are able to attain and maintain low body weight^16^, without reporting many of the hunger signals that are common among controls^14^, and cases frequently have comorbid diagnoses of gastric and digestive disorders^21,22^. Psychologically, individuals with AN experience undue fixation on weight or shape, are unable to appreciate the severity of their condition^16^, and often have co-existing psychiatric comorbidities such as depression, anxiety, obsessive compulsive disorder (OCD), and substance use^23–26^. AN genetic signatures are significantly correlated with metabolic biomarkers (insulin, cholesterol)^19^, general medical conditions (celiac disease, type I diabetes, peptic ulcer)^20^, and psychiatric symptoms (suicidality, depressive symptoms)^19,27^, and disorders (major depressive disorder, OCD)^19^. Thus, it is reasonable to hypothesize that polygenic risk for AN may encompass both metabolic and brain-based pathways, providing a useful test case to examine how pathway PRS approaches can yield insights into multi-systemic involvement in this disorder. This may also help us gain understanding into how polygenic risk is conferred to disease phenotype. Second, AN has historically been understudied relative to other psychiatric disorders, and new approaches to understanding the disorder are sorely needed^28^. AN has no effective or approved medications and bears one of the highest mortality rates among psychiatric disorders^29^, but it is also among the least in terms of the amount of research funding received^30^. Recent increases in sample sizes have yielded eight genome-wide significant loci for AN, demonstrating polygenicity at the variant level^19^. However, little is known regarding polygenicity at the pathway level. While past genetic studies of AN have suggested several pathways to be involved in the disorder development^19,31–33^, it is unclear how multiple pathways may work together to influence AN risk.

Through the approach of pathway PRS, which partitions genome-wide polygenic risk into pathways, we investigated the impact of multiple pathways simultaneously on disease development and comprehensively study polygenicity of AN at a higher biological level for the first time. We first examine how risk alleles within pathways impact likelihood of disease development and whether pathway PRS provided greater power to explain variance in AN than a traditional genome-wide PRS. Next, we asked whether pathway PRS could help explain the existence of cases with low genome-wide PRS, or controls with high genome-wide PRS. Could re-conceptualizing polygenic risk resolve these inconsistencies? To answer this, we explored pathway PRS residuals controlling for genome-wide PRS and examined whether individuals with low genome-wide PRS needed greater genetic liability from pathway PRS to become cases. We also examined whether aggregation, diversity and function of pathways matter, and whether diffused or concentrated risk across pathways is more likely to increase disease risk.

## Results

### AN genetic risk lies in specific pathways

Pathway PRS examines how genetic risk aggregates across the genome, focusing on biologically relevant gene sets and pathways. Application of pathway PRS to PGC-ED data (**Supplementary Table 1**) yielded pathway scores in 3,687 AN cases and 11,257 controls for 7,240 pathways in the Gene Ontology Biological Process (GOBP) and 421 pathways in our manual collection of psychiatric-related pathways^34^ (**Figure 1a**); in a univariate model, 497 pathway PRS was significantly associated with AN case/control status (applying a two-sided Bonferroni threshold for significance, p<6.5×10^−6^; **Supplementary Tables 2-3, Figure 1a**). In a joint model, 41 pathway PRS remained independently significant (P<0.05; **Supplementary Table 4, Figure 1b, Supplementary Fig. 1**), including pathway categories related to the nervous system, metabolic process, immune/response to stimulus, reproduction/developmental process, cellular process, cellular transport, and homeostatic process (**Table 1, Supplementary Table 5**). Pathway PRS were positively correlated with genome-wide AN PRS (mean *r*_pearson_ = 0.246, p<=3.38×10^−8^) such that individuals with high genome-wide AN genetic risk also had high pathway PRS regardless of case/control status (**Figure 1c**, Supplementary Results). In addition, high PRS controls had higher pathway scores than low PRS cases for 41 pathway PRS identified in the joint model (P < 0.05 for all but one pathway PRS; **Figure 1c**).

**Figure 1.**
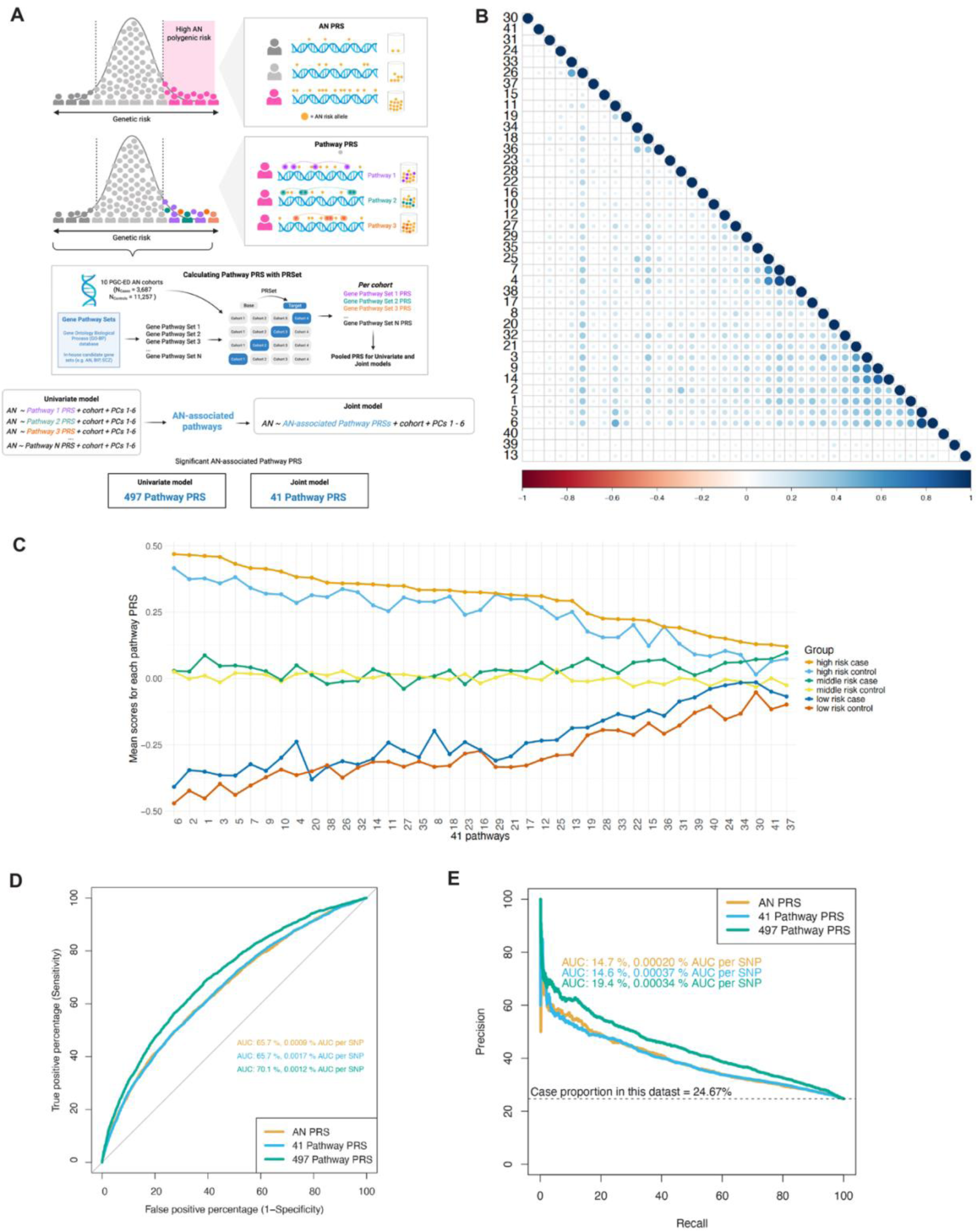
Identification of genetic pathways associated with anorexia nervosa. (A) A toy figure to illustrate the concept of pathway PRS and how we identified AN-related pathways. In contrast to the standard genome-wide PRS where all risk variants are counted, pathway PRS counts risk variants in each pathway, weighted by their effect sizes, for each individual. To identify pathways associated with AN, a total of 7,240 pathways from Gene Ontology Biological Process (GOBP) and 421 in-house collection of psychiatric-related pathways were examined. A total of 14,944 individuals in 10 cohorts from the Psychiatric Genomics Consortium Eating Disorders Working Group were included in this study. Pathway-based polygenetic risk scores (pathway PRS) were calculated using the PRSet function in PRSice-2. All pathway PRS calculated based on the leave-one-out analyses were then pooled into one dataset across 10 cohorts for the univariate and joint association analyses. A total of 497 pathway PRS were associated with AN in the univariate model after Bonferroni correction, of which 41 pathway PRS were also significantly associated with AN in the joint model, adjusting for cohorts and six genetic principal components. This figure was created using BioRender software (www.biorender.com). (B) Correlation plot of the 41 pathways PRS. The mean correlation across all 41 pathway PRS is 0.16, same as the mean correlation across 497 pathway PRS. See **Table 1** for the corresponding pathway names. (C) No single pathway PRS drives AN status. The mean pathway PRS for each of 41 pathway PRS is plotted, stratified by overall genetic risk (high, middle, low tertiles of AN PRS) and AN status (case/control). See **Table 1** for the corresponding pathway names. (D) Pathway PRS further explains AN risk compared to standard genome-wide PRS. The highest PR AUC was achieved by 497 pathway PRS, while the highest PR AUC per SNP was achieved by 41 pathway PRS. The number of SNPs included for AN PRS, 41 pathway PRS and 497 pathway PRS were estimated based on the largest included cohort (chop, **Supplementary Table 7**. The baseline line is y=24.67% as this equals the AN case proportion in the whole dataset. Here precision is defined as true positives over the sum of true positives and false positives, whereas recall (i.e., sensitivity) is defined as true positives over the sum of true positives and false negatives. A perfect prediction model would have a straight line at y=100% connected with x=100% at the top right corner. (E) The highest ROC AUC was achieved by 497 pathway PRS, while the highest ROC AUC per SNP was achieved by 41 pathway PRS. Here sensitivity (i.e., recall) is defined as true positives over the sum of true positives and false negatives, whereas specificity is defined as true negatives over the sum of true negatives and false positives. Therefore, 1 – specificity equals false positives over the sum of true negatives and false positives. A perfect prediction model would have a straight line at y=100% connected with x=0% at the top left corner. A random predictor with no discriminatory power would have an AUC of 50%.

**Table 1.** Effect size and p-value in the corresponding univariate models for 41 independent pathway PRS identified from the joint model. They are ordered from the most significant to the least based on their univariate p-value. These pathways are from either the Gene Ontology Biological Process (GOBP) in the Molecular Signatures Database (MSigDB version 7.4) or from our manually curated collection of psychiatric- related pathways^34^.

| # | Pathway | Coefficient | Standard Error | P-value |
| --- | --- | --- | --- | --- |
| 1 | Cell population proliferation [GOBP] | 0.20 | 0.02 | 1.13E-23 |
| 2 | Apoptotic process [GOBP] | 0.18 | 0.02 | 2.40E-19 |
| 3 | Positive regulation of protein metabolic process [GOBP] | 0.17 | 0.02 | 8.79E-19 |
| 4 | Cell cycle process [GOBP] | 0.16 | 0.02 | 3.85E-17 |
| 5 | Cell migration [GOBP] | 0.16 | 0.02 | 3.04E-16 |
| 6 | Locomotion [GOBP] | 0.16 | 0.02 | 5.66E-16 |
| 7 | Cell cycle [GOBP] | 0.15 | 0.02 | 3.10E-15 |
| 8 | Positive regulation of hydrolase activity [GOBP] | 0.15 | 0.02 | 9.24E-14 |
| 9 | Regulation of protein phosphorylation [GOBP] | 0.14 | 0.02 | 4.75E-13 |
| 10 | Schizophrenia (de novo) [manually curated] | 0.14 | 0.02 | 6.09E-13 |
| 11 | Cell part morphogenesis [GOBP] | 0.14 | 0.02 | 2.98E-12 |
| 12 | Exocytosis [GOBP] | 0.13 | 0.02 | 9.89E-12 |
| 13 | Schizophrenia (GWAS) [manually curated] | 0.13 | 0.02 | 1.41E-11 |
| 14 | Regulation of protein kinase activity [GOBP] | 0.13 | 0.02 | 1.36E-10 |
| 15 | Notch signaling pathway [GOBP] | 0.12 | 0.02 | 3.53E-10 |
| 16 | Lipid biosynthetic process [GOBP] | 0.12 | 0.02 | 3.62E-10 |
| 17 | Embryo development [GOBP] | 0.11 | 0.02 | 4.28E-09 |
| 18 | Cellular macromolecule catabolic process [GOBP] | 0.11 | 0.02 | 4.57E-09 |
| 19 | Cell-cell adhesion via plasma-membrane adhesion molecules [GOBP] | 0.11 | 0.02 | 9.88E-09 |
| 20 | Chemical homeostasis [GOBP] | 0.11 | 0.02 | 2.00E-08 |
| 21 | Positive regulation of intracellular signal transduction [GOBP] | 0.11 | 0.02 | 3.91E-08 |
| 22 | Cellular response to chemical stress [GOBP] | 0.11 | 0.02 | 4.71E-08 |
| 23 | Peripheral vasodilators [manually curated] | 0.11 | 0.02 | 4.78E-08 |
| 24 | Body mass index [manually curated] | 0.10 | 0.02 | 6.81E-08 |
| 25 | Negative regulation of cell cycle [GOBP] | 0.10 | 0.02 | 9.06E-08 |
| 26 | Response to abiotic stimulus [GOBP] | 0.10 | 0.02 | 9.52E-08 |
| 27 | Membrane organization [GOBP] | 0.10 | 0.02 | 1.03E-07 |
| 28 | Neuron apoptotic process [GOBP] | 0.10 | 0.02 | 1.03E-07 |
| 29 | Cellular protein-containing complex assembly [GOBP] | 0.10 | 0.02 | 1.30E-07 |
| 30 | Adenylate cyclase-inhibiting serotonin receptor signaling pathway [GOBP] | 0.10 | 0.02 | 1.56E-07 |
| 31 | tRNA metabolic process [GOBP] | 0.10 | 0.02 | 1.86E-07 |
| 32 | Response to cytokine [GOBP] | 0.10 | 0.02 | 2.66E-07 |
| 33 | Response to mechanical stimulus [GOBP] | 0.10 | 0.02 | 2.98E-07 |
| 34 | Anaphase-promoting complex dependent catabolic process [GOBP] | 0.10 | 0.02 | 3.39E-07 |
| 35 | Organelle assembly [GOBP] | 0.10 | 0.02 | 6.41E-07 |
| 36 | Regulation of mRNA metabolic process [GOBP] | 0.10 | 0.02 | 7.27E-07 |
| 37 | Positive regulation of release of cytochrome c from mitochondria [GOBP] | 0.09 | 0.02 | 1.14E-06 |
| 38 | Developmental process involved in reproduction [GOBP] | 0.10 | 0.02 | 1.23E-06 |
| 39 | Regulation of potassium ion transport [GOBP] | 0.09 | 0.02 | 1.93E-06 |
| 40 | Dicarboxylic acid metabolic process [GOBP] | 0.09 | 0.02 | 2.21E-06 |
| 41 | Neutral amino acid transport [GOBP] | 0.09 | 0.02 | 3.70E-06 |

Significance of pathway PRS was not driven by larger pathway size; on the contrary, pathway significance was negatively correlated with both pathway gene count (P = 2.90×10^−24^, *r* = −0.14) and variant count (P = 8.53×10^−33^, *r* = −0.17) (**Supplementary Fig. 2**), implying that smaller pathways were more likely to be associated with AN case/control status. Pathway PRS associations were also not driven by highly associated genes; p-value for the most significant gene within the pathway, calculated using MAGMA^19,35^, was moderately correlated with pathway significance in the univariate model (*r*_497pathway_ = 0.36, *r*_41pathway_ = 0.43, p<=0.0045), but not correlated with pathway significance in the joint model (P >=0.10, **Supplementary Fig. 3**).

### Pathway PRS explain more variance than genome-wide PRS

Modelling all 497 pathway PRS explained substantially more phenotypic variance (R^2^) in AN than the genome-wide PRS (9.20% vs. 3.91%, **Table 2**). Modelling the 41 independently associated pathway PRS performed similarly to genome-wide PRS (3.79% vs. 3.91%). Including genome-wide PRS and pathway PRS in the model had further significant improvement in predictive performance (10.65%, 5.70%, respectively, when modelling genome-wide AN PRS + (1) 497 pathway PRS or (2) 41 pathway PRS), suggesting that the pathway PRS do not simply recapitulate genome-wide signal (P_model_improvement_<=4.46×10^−9^). Permutation analyses selecting random pathway PRS from a list of 4,831 non-significant gene sets did not yield comparable R^2^ values, particularly for the 41 pathway PRS (R^2^ = 3.79% vs. mean R^2^_random_ = 0.42%, **Table 2**). Sensitivity analysis where random pathway PRS with matched pathway size were selected also showed that significant R^2^ by our identified pathway PRS was not due to chance or pathway size (Supplementary Results, **Supplementary Table 6**). Similarly, restricting the genome-wide PRS to include only the SNPs represented within the 41 pathways did not yield comparable R^2^ (R^2^_41pathway_ =4.64%, R^2^_AN PRS_ =4.66%, R^2^_AN PRS restricted to 41 pathway SNPs_ = 2.82%, **Supplementary Table 7**).

**Table 2.**
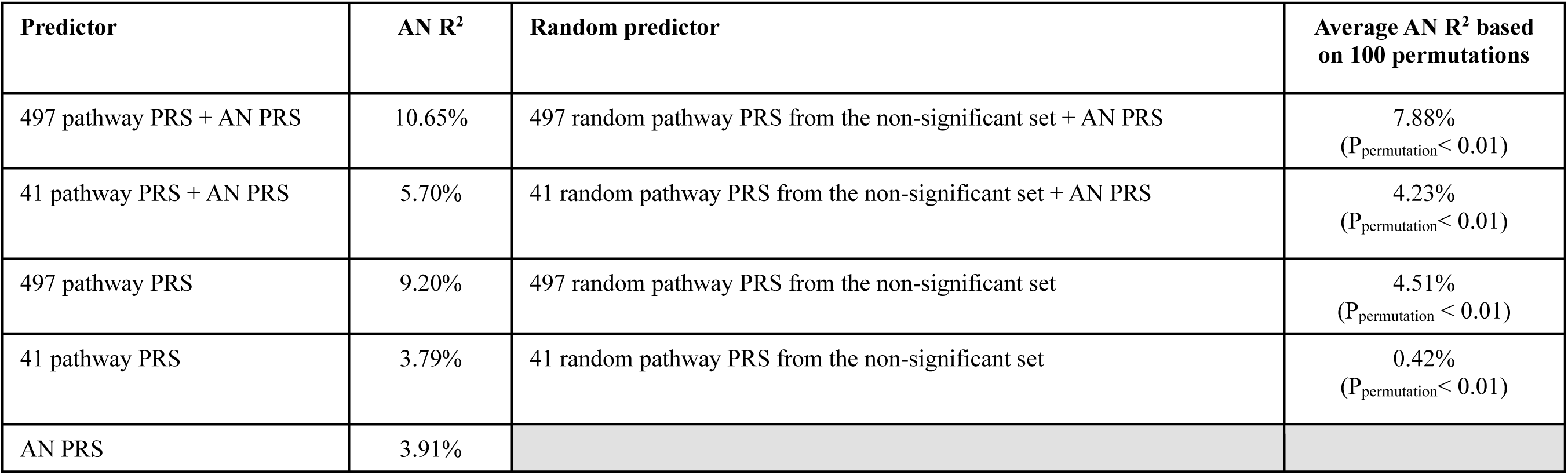
Variance in Anorexia Nervosa (R^2^) explained by different polygenic risk scores (PRS) Permutation was performed to examine the empirical significance of pathway PRS identified from the univariate and joint models in this study, using random pathways from the list of non- significant gene sets (i.e., 4,831 pathways with P>0.05 in the univariate model). The baseline model includes cohort and six genetic principal components. Abbreviations: AN=anorexia nervosa, AUC=area under the curve, BP=bipolar disorder, GOBP=Gene Ontology Biological Process, PC=principal component; PGC-ED=Psychiatric Genomics Consortium Eating Disorders Working Group, PR=precision-recall, PRS=polygenic risk score, ROC=receiver operating characteristic, SCZ=schizophrenia, SNP=single nucleotide polymorphism.

While PRS prediction is typically assessed in terms of phenotypic variance explained (R^2^), clinical utility and interpretability is more readily assessed through ability to predict case/control status. Therefore, we also assessed precision, recall (i.e., sensitivity), and specificity for genome-wide PRS, and our two sets of pathway PRS (497, 41 pathways), using positive predictive value (PPV, i.e., precision), precision-recall (PR) curves and Receiver Operating Characteristic (ROC) curves. Regardless of thresholds applied for case-control definition (Supplementary Results), overall precision was low, in line with expectations for polygenic prediction of psychiatric traits. Across the thresholds tested (Supplementary Results), PPV was highest when using the 497 pathway PRS model compared to 41 pathway PRS model or genome-wide AN PRS model (**Supplementary Tables 8-9;** PPV_497_=35.22%-44.46%). The 497-pathway model also had a greater area under the curve (AUC) for both the PR and ROC curves (**Figure 1d,e**). However, AUC per SNP was highest when considering the 41 pathway PRS model (**Figure 1d,e**). When genome-wide PRS and 497 pathway PRS were modeled together, precision, sensitivity and specificity all further improved but to a minor degree (PPV_497+AN-PRS_=35.63%-45.10%, **Supplementary Fig. 4, Supplementary Tables 10-11**).

### Pathway PRS partially explain how low-PRS cases develop disease

No single pathway PRS alone was sufficient to delineate cases from controls completely. Overall, genome-wide PRS was significantly correlated with risk in each pathway PRS, and no pathway PRS was consistently higher in all cases compared to all controls (**Figure 1c)**. Rather, when considering all PGC-ED samples in our study, we found that controls with high genome-wide PRS had higher pathway PRS than low genome-wide PRS cases (**Figure 1c**).

We hypothesized that perhaps the combination of genome-wide PRS and pathway-specific PRS has to meet some unknown threshold in order for an individual to develop disease. Thus, we propose that individuals with lower genome-wide PRS will have higher pathway-PRS, in order to meet this threshold. (**Figure 2a**). Individuals with high genome-wide PRS conversely have a lower required pathway-specific threshold to develop a disorder.

**Figure 2.**
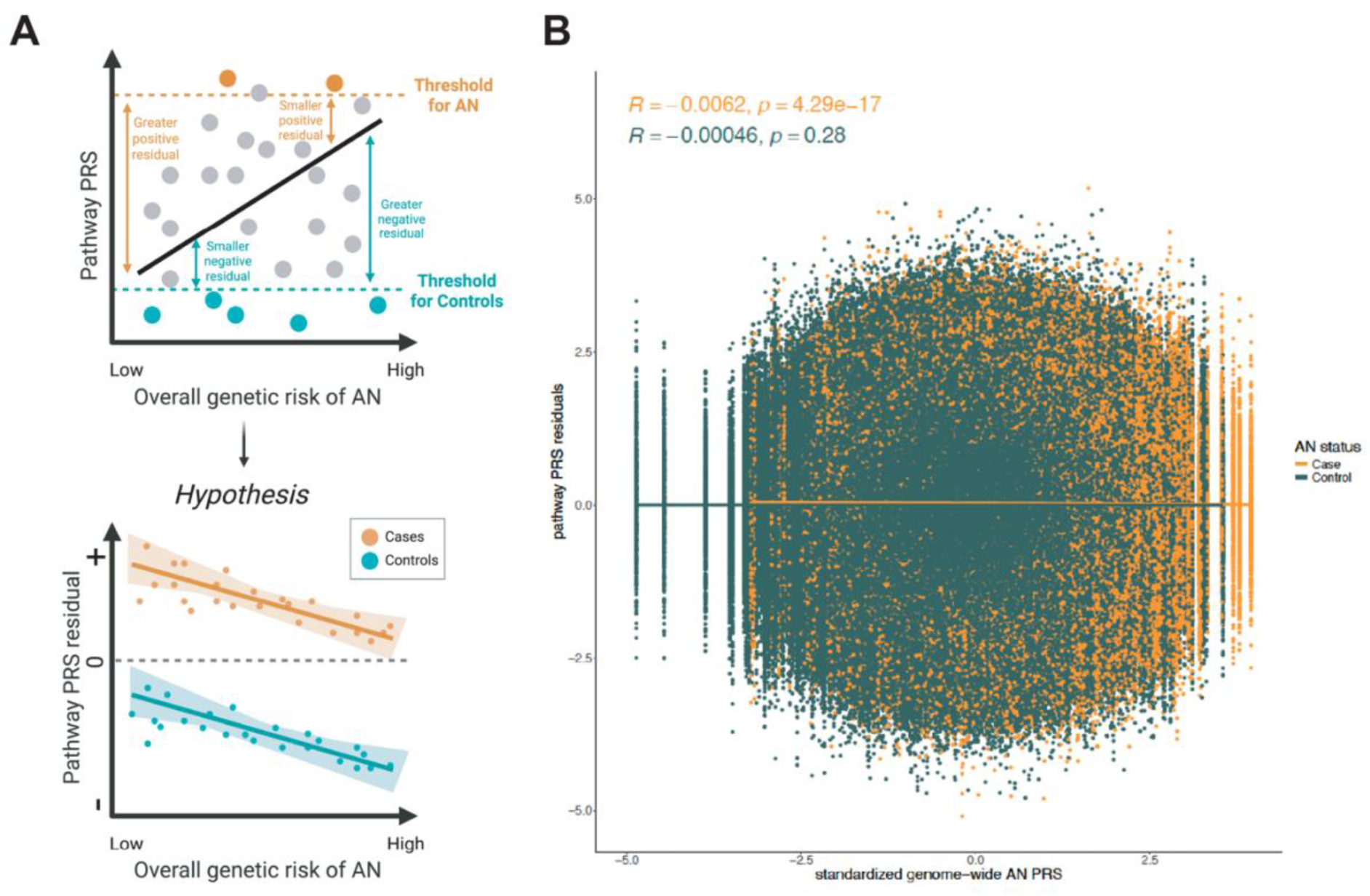
Pathway PRS partially explains why individuals with low genetic risk could develop AN. (A) A toy figure to illustrate our hypothesis of genetic liability threshold such that among cases, individual with a lower AN PRS would have higher pathway PRS residuals (i.e., greater positive residuals) to be a case, whereas among controls, individuals with a higher AN PRS would have lower pathway PRS residuals (i.e., greater negative residuals) to be a control. Our finding suggests that our hypothesis for cases is true, but not for controls. This figure was created using BioRender software (www.biorender.com). (B) Correlation between genome-wide AN PRS (x axis) and pathway PRS residuals (y axis) for 497 significant pathways identified from the univariate models. Pearson correlation and its associated p-value are shown for both cases and controls. While we observed negative correlations between pathway PRS residuals and AN PRS for both cases and controls as we hypothesized, only the correlation in cases was significant, suggesting pathway PRS contributes to the genetic liability threshold to become a case for those with lower AN PRS. Abbreviations: AN=anorexia nervosa, PRS=polygenic risk score.

To probe this hypothesis, we calculated pathway PRS residuals, adjusting each pathway score for genome-wide PRS scores (**Supplementary Fig. 5**). We found that cases in general had positive pathway residuals (**Supplementary Fig. 5**) and that AN cases with lower genome-wide PRS tended to have higher pathway PRS residuals compared to cases with higher genome-wide PRS (P = 4.29×10^−17^, **Figure 2b**).

### Combination of pathways matters: specific pathway PRS pairs confer higher risk

To examine whether specific combinations of pairs of pathways within four biological groups (brain, metabolism, immune/stress, and reproduction) confer higher or lower risk, we calculated case proportions among all individuals who had top pathway PRS scores (defined as top 10% pathway PRS, Top_10%_) across any two pathways (**Supplementary Tables 12-15**). AN percentages were highest (>40%) among individuals with Top_10%_ risk for 86 specific pathway pairs (i.e., top metabolic-metabolic pathway pair: *glycosyl compound biosynthetic proce*ss and *negative regulation of nucleobase-containing compound metabolic process* [43.75% AN, *r*_pathway_pair_ = 0.042], top brain-metabolic pathway pair: *regulation of NMDA receptor activity* and *carbohydrate derivative metabolic process* [43.24% AN, *r*_pathway_pair_ = 0.11], and top brain-brain pathway pair: *glutamatergic synapse* and *axon development* [42.21% AN, *r*_pathway_pair_ = 0.034], **Supplementary Figs. 6-7, Supplementary Table 16**). Case proportions were lowest (19.08% AN, *r*_pathway_pair_ = 0.043) among those with high scores in *alpha-amino acid metabolic process* and *protein dephosphorylation* (P_Bonferroni_ = 0.031 compared to the pathway pair with the highest AN%, **Supplementary Table 16**).

### Number of pathways matters: AN risk accumulates across relevant pathways

Our results show that higher scores of each of the 497 pathway PRS were associated with case status. Next, we sought to understand how accumulation of these pathway PRS confer risk. We also asked whether distribution of risk across pathways matters, focusing on the number, diversity, and functional consequences of pathways.

First, we examined whether individuals who had high genetic risk (i.e., Top_10%_ pathway PRS) across multiple pathways had a higher risk of disease (**Figure 3a**). Across all cohorts, individuals with high scores across multiple pathways were more likely to be cases (Pearson correlation between the number of Top_10%_ pathways and case/control ratio: 497 pathways: *r* = 0.74, p = 7.98×10^−21^, **Figure 3b**; 41 pathways: *r* = 0.74, p = 4.0×10^−4^, **Figure 3c**; compared to genome-wide AN PRS deciles: *r* = 0.96, p = 1.56×10^−5^, **Figure 3f**); permutation tests indicate that this is not due to chance (P_497pathway_<0.0001, P_41pathway_=0.036 based on 10,000 permutations, **Supplementary Fig. 8**), and repetition across other thresholds (i.e., Top_5%_, Top_20%_, Top_30%_ and Top_50%_) yields the same positive and significant relationship (P<=9.6×10^−14^, **Supplementary Fig. 9**). To ensure this effect is not completely due to correlation with the genome-wide AN PRS, we repeated our analysis removing all individuals in the top 10%, 30%, 50%, 70% and 80% of AN PRS. A significantly positive correlation persisted even among individuals at the bottom 30% of AN PRS (N = 3,342), but was no longer significant when top 80% samples were removed (N = 1,782, **Supplementary Figs. 10-11**). Further, a significantly positive correlation between AN proportion and Top_10%_ pathway count also persisted for 497 pathways after adjusting for genome-wide PRS (P=8.88×10^−6^, **Figure 3d**), but no longer significant for 41 pathways (P=0.37, **Figure 3e**). The converse relationship is also true: individuals with low scores across multiple pathways were significantly less likely to be cases (correlation between the number of Bottom_10%_ pathways and case/control ratio among 497 pathways: *r* = −0.64, p = 3.7×10^−14^, **Supplementary Fig. 9**).

**Figure 3.**
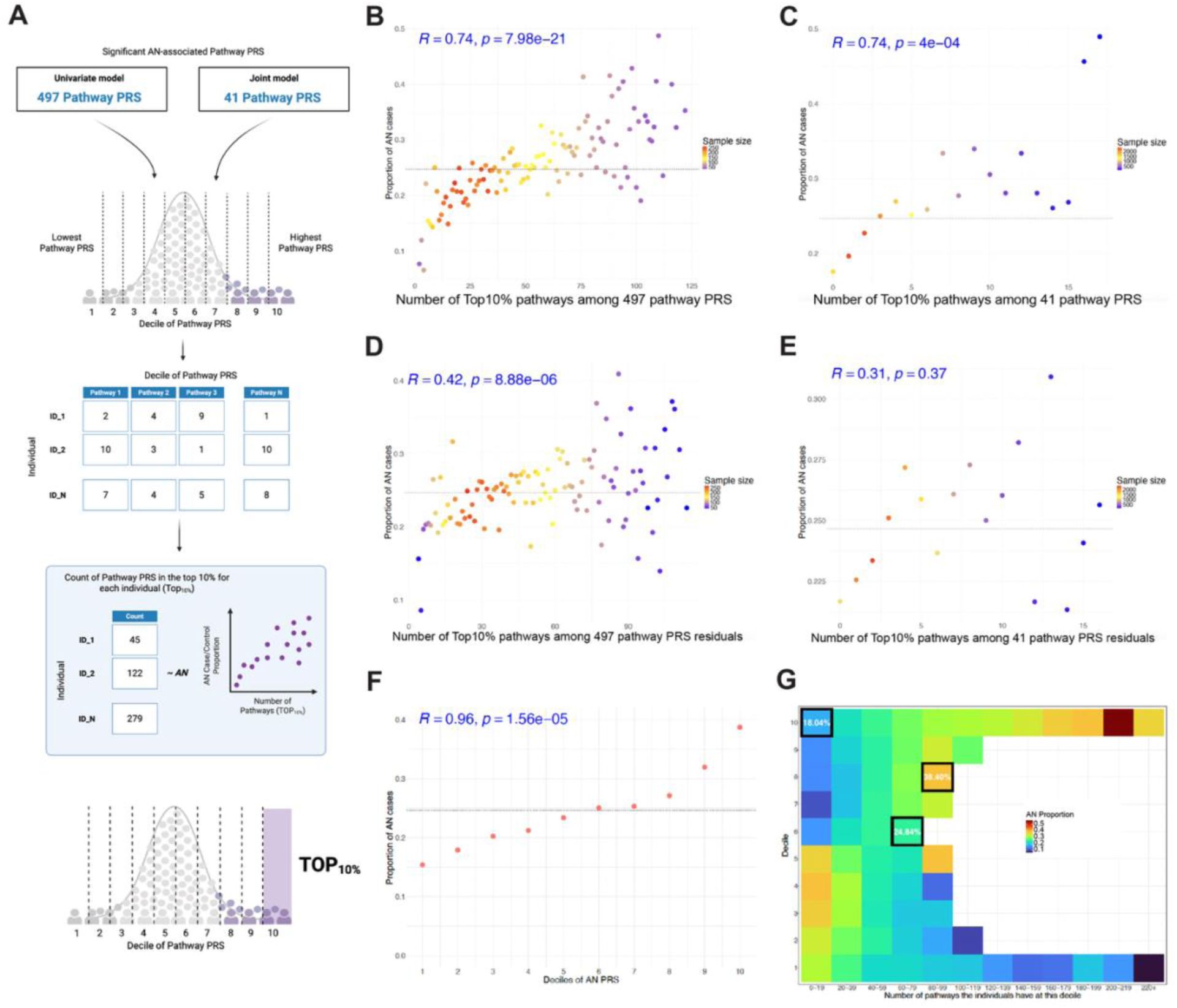
Genetic risk in more pathways confers higher AN risk. (A) A toy figure to illustrate how we calculate top pathways. We first converted pathway PRS into deciles and then counted the number of pathway PRS in the top 10% (i.e., decile 10) for each individual. Correlation was then examined between Top_10%_ pathway count and AN case proportion. This figure was created using BioRender software (www.biorender.com). (B) 497 pathways: Correlation between number of pathways with top 10% risk (x axis) and AN case proportion (y axis) for 497 significant pathways identified from the univariate models. Pearson correlation and its associated p-value are shown in blue. The baseline AN proportion in the dataset is plotted as a dotted line (y=24.67%). (C) 41 pathways: Correlation between number of pathways with top 10% risk (x axis) and AN case proportion (y axis) for 41 significant pathways identified from the joint model. (D) 497 pathway residuals: Correlation between number of pathways with top 10% risk (x axis) and AN case proportion (y axis) for residuals of 497 significant pathways adjusted for AN PRS, cohort, and 6 genetic PCs. (E) 41 pathway residuals: Correlation between number of pathways with top 10% risk (x axis) and AN case proportion (y axis) for residuals of 41 significant pathways adjusted for AN PRS, cohort, and 6 genetic PCs. (F) Relationship between AN PRS decile (x axis) and AN case proportion (y axis) is plotted to serve as a positive control and reference for pathway PRS in terms of the correlation strength with AN risk. (G) AN case proportion is plotted for individuals that fell in certain pathway risk level for certain numbers of pathways. We first calculated the number of pathways each individual had at each decile. We then summed all the individuals at certain decile for certain number of pathways and calculated how many AN cases were present in that category. Overall, individuals with more pathways at top 10% had greater AN risk, and those with more pathways at bottom 10% had lower AN risk. Through this, we can also examine concentrated versus diffused genetic risks. For example, (1) individuals at the top decile for 0-19 pathways had 18.04% AN, (2) individuals at the top 5^th^ decile for 60-79 pathways had 24.84% AN, and (3) individuals at the top 3^rd^ decile for 80-99 pathways had 38.40% AN. Individuals in each tile are not mutually exclusive from each other. **Supplementary Figure 12** has more details on the extent of individual overlap for different tiles. Tiles with a total head count less than 10 are not plotted. Abbreviations: AN=anorexia nervosa, PC=principal component, PRS=polygenic risk score.

Next, we used the same design to test whether polygenicity confers higher risk when having top genetic risks concentrated within a small number of pathways, or when having above-average risks spread across many pathways; that is, whether concentrated or diffused genetic risk throughout the genome is more likely to lead to disease. To test this, we compared how AN case proportions change according to the number of pathways at the top 1^st^, 3^rd^, and 5^th^ deciles. We found that individuals who had top 10% genetic risk in a few pathways had lower AN case proportion (i.e., top 10% in 0-19 pathways [18.0% AN, N=3,170]) compared to those with top 3^rd^ or 5^th^ decile genetic risk in more pathways (i.e., P = 5.4×10^−15^ against individuals with pathway PRS in 80-99 pathways at top 3^rd^ decile [38.4% AN, N=250]; P = 8.2×10^−10^ against individuals with pathway PRS in 60-79 pathways at top 5^th^ decile [24.8% AN, N=2,355], **Figures 3g**, **Supplementary Figs. 12-13**).

### Type of pathway matters: more diverse pathways confer higher risk

We used two complementary approaches to test whether risk concentrated in more or less similar pathways confers higher risk for disease development; first, comparing correlation between pathway PRS scores and case/control likelihood; second, comparing Jaccard Index scores across all Top_10%_ pathways for cases *versus* controls. These analyses yield consistent results. First, a higher correlation between pathway PRS is negatively correlated with AN risk (P_497pathway_=1.42×10^−40^, P_41pathway_=0.0032, **Supplementary Fig. 14**), suggesting that a more diverse set of pathways confers higher risk of AN. Second, we found that AN cases had significantly lower Jaccard index scores (i.e., less similar) compared to controls (P = 1.09×10^−35^, **Supplementary Fig. 15**). This effect persisted when controlling for the overall AN PRS (P = 2.45×10^−4^).

### Pathway function matters: brain and metabolic pathways confer higher risk

To test whether the biological grouping of pathways had an impact on AN risk (**Figure 4a**), we repeated our previous case/control proportion analysis partitioning our 497 pathways into brain, metabolic, immune/stress, and reproduction-related pathways (**Supplementary Tables 12-15**). There is a significant enrichment of AN cases among individuals with more Top_10%_ brain (*r* = 0.88, P = 9.97×10^−8^), metabolic (*r* = 0.71, P = 3.90×10^−8^) and immune/stress-related pathways (*r* = 0.78, P = 2.89×10^−5^), but not reproduction-related pathways (*r* = 0.16, P = 0.69) (**Figure 4b-e**). Our sensitivity analysis indicates that this is not driven by pathway numbers (Supplementary Results). We also observed a combinatorial effect, whereby individuals with higher numbers of both brain and metabolic pathways were more likely to be cases (e.g., 40.63% AN for those with 8 brain pathways and 13 metabolic pathways, compared to 10% AN for those with 0 brain pathway and 0 metabolic pathway at top 10%, P_Bonferroni_ = 0.0033, **Figure 4g**).

**Figure 4.**
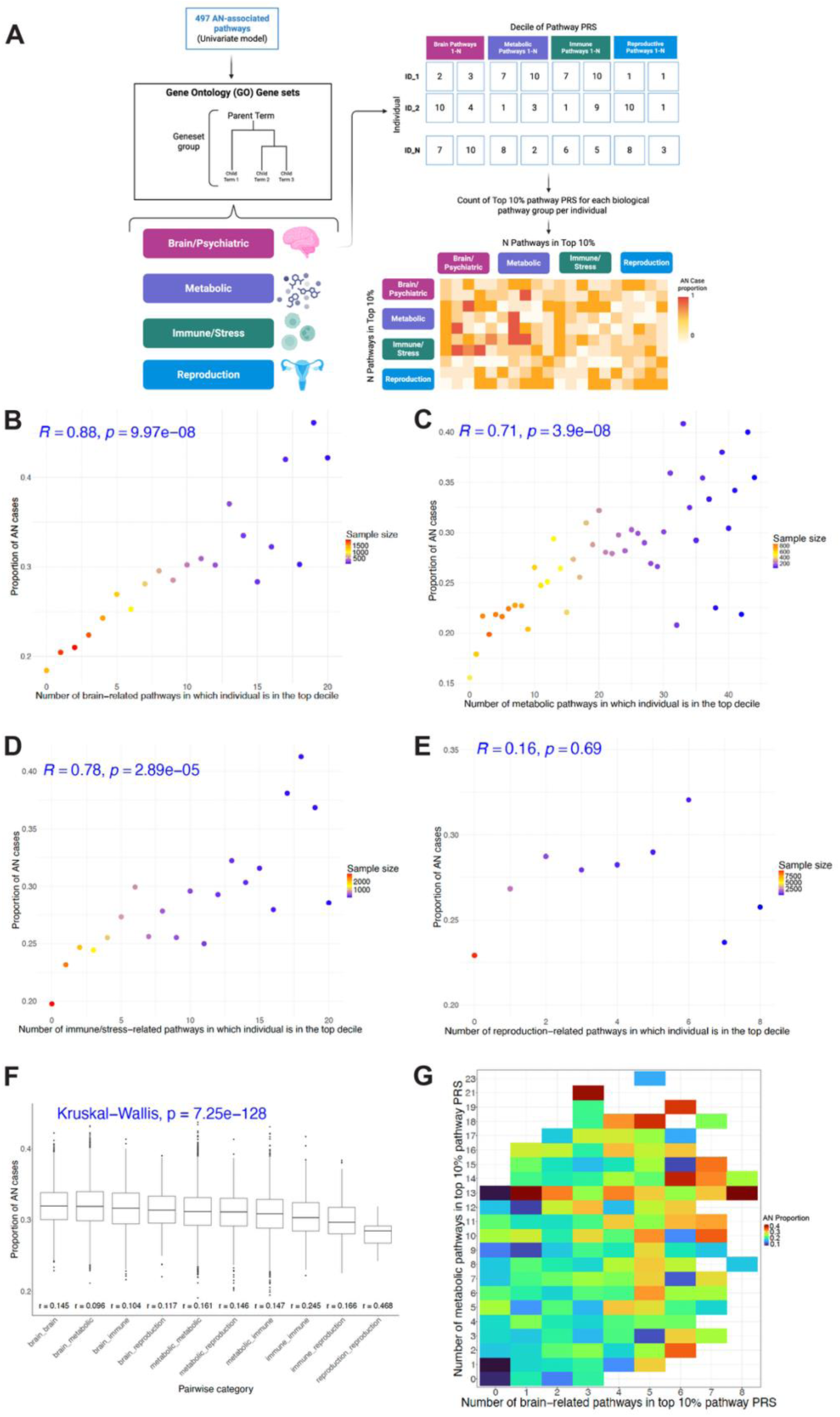
Pathway function influences AN risk. (A) A toy figure to illustrate how pathway count in each of the four biological groups (i.e., brain, metabolic, immune/stress, reproduction) was generated for each individual. Four biological groups were identified based on the names of the 497 significant pathways themselves or their associated parent nodes given the tree structure of Gene Ontology using the GOfuncR package in R. We classified a total of 49 brain-related pathways, 126 metabolic pathways, 39 immune/stress-related pathways, and 9 reproduction-related pathways. Each pathway pair includes individuals whose pathway PRS were ranked at top 10% for both pathways. AN case proportion was then calculated for each within-function and across-function pathway pairs. The exact AN case proportion for each pathway pair is included in **Supplementary Table 16**. This figure was created using BioRender software (www.biorender.com). (B) Number of pathways where the individual ranks in the top 10% for pathway PRS in 49 brain-related pathways. Each dot represents a group of individuals with certain number of Top_10%_ pathways. Only dots with more than 30 individuals are included for plotting. Pearson correlation and its associated p-value are shown in blue. (C) Number of pathways where the individual ranks in the top 10% for pathway PRS in 126 metabolic pathways. (D) Number of pathways where the individual ranks in the top 10% for pathway PRS in 39 immune/stress-related pathways. (E) Number of pathways where the individual ranks in the top 10% for pathway PRS in 9 reproduction-related pathways. (F) AN proportion distribution for pairwise pathways within and across functions. Among 497 AN-associated pathways from univariate models, 223 pathways were classified into these 4 groups and were used for plotting here. The order is based on median AN proportion per category. Significance of overall difference across all groups was examined by Kruskal-Wallis test. P-values for all pairwise comparisons across 10 categories are included in **Supplemental Table 17**. The mean correlation for each group is shown at the bottom. (G) AN proportion was shown in a heatmap format for individuals with certain number of brain-related pathways (x axis) and certain number of metabolic pathways (y axis) where they rank at top 10%. Abbreviations: AN=anorexia nervosa, PRS=polygenic risk score.

To study whether cross-function or within-function pathways have more effect on AN risk, we examined the relationship between AN proportion and ten types of pathway pairs, allowing within-function and cross-function comparisons (e.g. brain-brain, metabolic-metabolic, brain-metabolic, et cetera, **Figure 4f, Supplementary Fig. 16**). Individuals with brain-brain pathway pairs and brain-metabolic pathway pairs had the highest AN risk (P_difference between them_ = 0.75, P_difference from other types_<=0.013) (**Figure 4f, Supplementary Table 17**); in general, case proportions were highest in pairs of pathways including brain, then metabolic, then immune, then reproductive (**Figure 4f**). These patterns were generally consistent among the smaller set of 41 pathways (**Supplementary Fig. 16**). AN risk was negatively correlated with pathway similarity among within-function groups and among cross-function groups, as in the full sample.

## Discussion

Our study examined how genetic risk for AN accumulates across pathways, providing insights into the polygenic nature of AN. We were particularly motivated to examine the genetic risk underpinning AN, given the multi-systemic nature of the disease, with demonstrated metabolic and psychiatric involvement. Our results reinforce our previous findings of metabo-psychiatric underpinnings; polygenic risk that accumulates in brain- or metabolic-pathways confers the greatest risk for AN, and risk is heightened if both brain and metabolic pathways are impacted. Moreover, our results identify specific pairs of pathways that may confer especially high AN risk; we identified 86 pathway pairs for which individuals with Top_10%_ PRS had almost double the risk of developing AN (i.e., AN%>40%) compared to the whole cohort (AN% = 24.67%). In addition, we found that Top_10%_ pathways were more diverse (i.e., less similar to each other) in AN cases compared to controls. Our analyses represent a major advance in our understanding of how polygenic risk accumulates for development of AN.

On a broader scale, our results provide insights into the nature of polygenicity in general. We examine how polygenic risk may be partitioned into pathways, and how risk accumulates across and within those pathways to answer major questions about how polygenicity occurs at a higher biological level. For example, we show that the aggregation of risks across pathways matters. We observed a robust positive relationship between AN risk and high scores (Top_10%_) in more pathways. This positive relationship persisted both within functions (e.g., brain, metabolic) and across functions (e.g., brain + metabolic). The positive relationship between AN and pathway count also persisted regardless of the overall genetic risk level (e.g., when removing individuals with top 70% AN PRS) or the threshold used to define top pathways (e.g., Top_5%_ to Top_50%_).

Although at the population level, disease risk may be predicted by aggregation of risk alleles weighted by effect size (i.e., a classical PRS), more nuanced insights may be obtained by considering the functional units within which the variant is located (here, genes and pathways). We show that specific pathways confer higher risk than others, and that the significance of these pathways is not driven by pathway or gene size (**Supplementary Fig. 2**), implying that polygenicity is not random across the genome.

On the basis of polygenicity, we also examined whether individuals with risk in the top decile in a small number of pathways (i.e., concentrated risk) or individuals with lower risks (top 3^rd^ and 5^th^ deciles) across many pathways (i.e., diffused risk) had a higher disease risk. While we found that diffused risk was associated with higher AN proportion (**Figure 3g**), we noted that the reduced AN risk for individuals with high genetic risk in only a few pathways could also be because these individuals had bottom 10% of genetic risk in many other pathways (**Supplementary Fig. 12**a), possibly including some pathways that are necessary for disease development. In addition, those with top 3^rd^ or 5^th^ decile genetic risk in more pathways also had many pathways at the top 10% genetic risk (**Supplementary Fig. 12**b,c). Therefore, it is impossible to compare individuals with diffused risk only versus those with concentrated risk only. Instead, we examined these individuals from a holistic perspective based on their pathway risk distribution (**Supplementary Fig. 12**).

Next, we examined the pathways themselves and asked whether any features of these pathways confer higher risk. We found that pathway diversity and function both matter: more diverse pathways were associated with a greater risk of disease (**Supplementary Figs. 14-15**), and high scores in multiple psychiatric and metabolic pathways were associated with greater disease risk (**Figure 4g)**. Further, while previous literature has suggested several pathways to be involved in AN etiology, including those related to the brain^31,36^, immune system^33^, reproduction/development^19^, and metabolism^20^, it remains unclear whether these pathway functions are equally important or some have a greater impact than others. Through evaluating within-function and across-function pathway pairs, our results suggested that on average brain-related pathways were associated with the highest AN risk, followed by metabolic pathways and then immune/stress-related pathways (**Figure 4f, Supplementary Fig. 16**). While we also found several significant pathways related to reproduction (e.g., developmental process involved in reproduction, embryo development), aggregation of reproduction-related pathways (**Supplementary Table 15**) was not significantly associated with AN risk (**Figure 4e**). It is possible that the association may change if we expanded our reproduction-related pathways to all development-related pathways. However, we decided to keep reproduction-related pathways for examination considering that patients with AN may be at a higher risk of reproductive issues^37^, and it is worth exploring whether there is any genetic link between reproduction-related pathways and AN. In addition, a general development theme is too broad and have multiple overlap with pathways included in other categories (e.g., central nervous system development in the brain category and immune system development in the immune/stress-related category).

Further, we observed a general trend that AN risk was lower when correlation between pathway pairs was higher. This applies to within-function pathway pairs, across-function pathway pairs (**Figure 4f**), and all pathway pairs across the 497 pathways (**Supplementary Fig. 14**). Given this, the fact that within-function pathway pairs had similar or even higher AN risk compared to their cross-function counterparts (e.g., brain-brain vs. brain-metabolic; metabolic-metabolic vs. metabolic-reproduction; immune-immune vs. immune-reproduction) while having a higher mean correlation (**Figure 4f**) suggest that there may exist some synergistic effect on disease risk between pathways within the same functional group to partially offset the higher correlation between them.

On a population level, one ongoing question in polygenic analyses of complex traits is the existence of cases with low PRS, or controls with high PRS. To some extent, these instances may be explained by the unaccounted-for role of rare or protective variants, environmental and societal factors, or under-diagnosis among controls. However, it is also possible that polygenic risk scores aggregate meaningful genetic variation together with irrelevant variation; essentially obscuring case/control stratification through signal:noise ratio. We hypothesized that accounting for patterns among risk alleles may resolve this mismatch between genome-wide PRS and trait outcome. By partitioning risk into biologically relevant pathways, we hoped to examine whether ‘low-risk’ cases in fact have substantial risk concentrated in relevant pathways, while ‘high-risk controls’ have risk allele scattered at random throughout the genome.

Specifically, we hypothesized that a combination of genome-wide and pathway-specific genetic risks are required to meet a certain threshold for disorder development. Thus, individuals with low overall PRS would require higher pathway PRS residuals to develop a disorder (**Figure 2**), and individuals with high genome-wide PRS conversely have a lower required threshold for pathway PRS to develop a disorder. Indeed, our results support this hypothesis: cases with low genome-wide PRS had higher pathway PRS residual scores, compared to cases with high genome-wide PRS, implying that pathway PRS may help to resolve low PRS cases. However, our data do not support a similar relationship for controls; while high-risk controls tended to have more negative pathway residual scores than low-risk controls, this relationship was not significant, implying that pathway PRS may help to understand why individuals with low genetic risk become a case, but it may not help address why those with high genetic risk remain to be disease-free. This may reflect the unaccounted environmental risk factors or triggers, precluding disorder development in these high-risk individuals. Alternatively, we consider that the pathways identified in this study were selected as conferring risk for AN, such that high scores are associated with increased risk. We did not set out to identify protective or resilient PRS by comparing high-risk controls against all cases. Further, it is not necessarily true that pathways must confer both increased and decreased risk: rather, it is likely often the case that high scores may increase risk, but low scores may confer only average risk for disease, rather than acting as protective.

We also examined the potential in clinical utility of pathway PRS by examining its performance of disease prediction. While the precision in AN prediction still remains low regardless of using either genome-wide PRS or pathway PRS, polygenic risk accumulated within these pathways significantly increased the amount of phenotypic variance explained compared to standard genome-wide PRS (**Table 2**), and improved ability to distinguish cases from controls (**Figure 1d,e, Supplementary Fig. 4, Supplementary Tables 8-11**). This improvement by pathway PRS has also been observed when being used to predict other health outcomes, including psychosis^38^ and treatment response to lithium in patients with bipolar disorder^39^.

We note some limitations of our study. First, our analysis focuses on germline genetic variation, rather than incorporating biological consequences. Analyses that integrate polygenic risk with functional data (for example, expression quantitative trait loci, cell-type-specific genes) may yield more actionable insights^40–42^. Second, our analysis focuses specifically on risk-increasing pathways, rather than protective pathways or those conferring resilience, potentially limiting insights that we obtain with regards to controls. Third, these AN-associated pathways were identified based on individuals with European ancestry. It is yet to be explored if the same biological pathways are shared across ancestral groups. However, current genetic studies of AN are mostly based on participants with European ancestry and cross-ancestry analysis would be possible in the future as more genetic data become available^28^. Fourth, AN is a heterogeneous trait, with distinct disease subtypes (restrictive, binge-purge). As it stands, not all cases in PGC-ED have available subtype information, and data with available subtypes and raw genotype data are too small to analyze in these studies. However, it is likely that different genetic etiologies that underlie these subtypes may be reflected by different pathways identified in this study. Future analyses with access to raw data from additional cases and controls should examine whether specific patterns of pathway PRS confer risk for different subtypes or indeed specific behaviors.

A reasonable concern about pathway PRS is that these results may be driven by common confounders such as pathway size, the presence of significant genes, SNP-gene assignment approaches, or overfitting within cohorts. We guard against these in our study, verifying that neither SNP nor gene count positively correlated with pathway significance. In fact, among significant pathways, those with smaller SNP count or gene count tended to have higher significance, indicating that smaller pathways with a more defined role reflect genetic etiology of AN better than larger pathway PRS, possibly due to a higher signal:noise ratio in smaller pathways. P-values of the most significant gene also did not highly correlate with pathway-level associations. Moreover, many significant pathways did not include genes reaching Bonferroni significance. For example, while the adenylate cyclase-inhibiting serotonin receptor signaling pathway reached Bonferroni significance in our analysis (P_univariate_ = 1.56×10^−7^, P_joint_ = 0.00014), only 2 out of 7 genes in this pathway reached nominal significance (P*_HTR1B_* = 0.0016, P*_HTR1E_* = 0.0010), suggesting a collective effect of this pathway on AN status rather than being entirely driven by a few significant genes. We also performed analysis to ascertain appropriate gene flanking windows for SNP assignment (**Supplementary Fig. 17**) and further demonstrated that the pathway PRS approach has higher cross-cohort replicability (**Supplementary Fig. 18**), with substantially higher correlations in effect sizes between cohorts compared to MAGMA^35^, a commonly used tool for gene set analysis (Supplementary Results, **Supplementary Figs. 19-21**).

In summary, our study demonstrates polygenicity at the pathway level, evidenced by increased AN risk when pathway risks aggregated within and across functions. The distribution of risk alleles is not random and accumulates into biologically relevant pathways, with brain-related pathways having the largest impact on AN risk, followed by metabolic pathways. At a more nuanced level, specificity of pathway pairs matters as some convey more than double the risk than others. While we cannot completely resolve genetic risk on an individual level, we found that pathway PRS contributed to disease development in low-risk individuals. As the promise of precision medicine requires translation of PRS from a global (i.e., population level estimate) towards an individual level estimate, our findings highlight a translational potential of pathway- based PRS to prioritize pathways for functional investigation and to improve disease prediction and subtype identification in the future.

## Supporting information

Supplementary results and figures

## Online Methods

### Participants

A total of 3,687 anorexia nervosa (AN) cases and 11,257 controls from ten cohorts (N_total_=14,944, AN case proportion = 24.67%) in the Psychiatric Genomics Consortium Eating Disorders Working Group (PGC-ED) were included in this study^19^. The diagnosis of AN was established based on standard criteria (i.e., Diagnostic and Statistical Manual of Mental Disorders [DSM-III-R, DSM-IV] and International Classification of Diseases [ICD-9, ICD-10]) using structured clinical interviews, questionnaires, or hospital/register records. The controls largely came from unscreened cohorts^19,43–47^. But given the low population prevalence of AN^48^, the inclusion of a small number of AN cases in controls should have minimal impact^20^. All study participants were of European ancestry. Strict quality control was applied to ensure quality of genetic variants and study samples and to remove ancestry outliers and familial relatedness using the PGC Rapid Imputation For Consortias Pipeline (Ricopili)^49^. Duplicated SNPs were removed and only SNPs with < 1% missing rate and > 5% MAF were included for calculation of polygenic risk scores (PRS). The final numbers of genetic variants, cases, and controls per cohort were included in **Supplementary Table 1**. Further details about the cohort, case ascertainment, genotyping platform, and quality control were described elsewhere^19^. Ethical approval and participant consent were obtained for each contributing cohort^19^.

### Calculation of pathway PRS

Pathway PRS is a weighted sum of genetic risk variants within a biological pathway, where the weight is the odds ratio of the variant on the outcome of interest (i.e., anorexia nervosa). In this study, individual-level pathway PRS were calculated using the PRSet feature^50^ in PRSice-2 (v2.3.1)^51^. We employed a leave-one-out approach, using genome-wide summary statistics of AN^19^ from all but one cohort to serve as the ‘base’, to calculate pathway PRS in the remaining ‘target’ cohort. This process was repeated 10 times to calculate pathway PRS in all ten cohorts (**Figure 1a**). An *a priori* set of 7,240 pathways from Gene Ontology Biological Process (GOBP) (Molecular Signatures Database [MSigDB] version 7.4)^52–54^ and a manual in-house collection of 421 psychiatric-related pathways^34^ were used to define genes included in each pathway. The genome boundary of each gene was defined based on an ENSEMBL^55^ (GRCh37.75) gene transfer format (GTF) file suggested by PRSet^50^. We included variants that were 35 kilobases (kb) upstream and 10 kilobases (kb) downstream of each gene in the calculation of pathway PRS^50^ given genetic regulatory elements often reside in the gene flanking region. We also conducted sensitivity analysis on the size of flanking region and evaluated performance of 35 kb upstream/ 10 kb downstream versus 1 megabase (mb) upstream / 1 mb downstream, reflecting standard assumptions about gene cis regions. Our results suggested a higher replication rate of pathways across cohorts using the 35 kb/ 10 kb cutoff (**Supplementary Fig. 17**) and therefore we used this default setting of PRSet for calculation of pathway PRS (--wind-3 10kb, --wind-5 35kb). Any single nucleotide polymorphism (SNP) variant that has a linkage disequilibrium (LD) correlation of 0.8 or greater with variants on or near the genes was also included (--proxy 0.8). To keep independent genetic markers in PRS calculation, SNPs were clumped within each pathway using the default of 1 mb size for PRSet (i.e., clumping was performed for a window size of 2mb with the index SNP in the middle, --clump-kb 1mb) with filter thresholds of R^2^ of 0.1 (i.e., SNP pairs with LD correlation greater than 0.1 were clumped and the SNP with a more significant p-value was kept, --clump-r2 0.1) and p-value of 1 (i.e., no SNP filter was performed based on p-value for clumping, --clump-p 1). For the calculation of pathway PRS, p-value thresholding was set at 1 as default by PRSet^50^. After pathway PRS were calculated for the 10 cohorts, they were pooled into one dataset.

### Identification of AN-related pathway PRS

Logistic regression model was performed where AN status was the outcome and a pathway PRS was the predictor variable, while adjusting for cohorts and 6 genotype-derived principal components (AN ∼ PRS_pathway_ + cohort + PC1 + … + PC6). AN-associated pathways were identified if they passed the stringent Bonferroni-corrected significance (P < 6.5×10^−6^). Next, to identify independent pathways, we ran a joint model which included all Bonferroni-significant pathway PRS from the univariate model, adjusting for cohort and 6 PCs (AN ∼ PRS_pathway_1_ + … + PRS_pathway_497_ + cohort + PC1 + … + PC6). Independent AN-associated pathways were identified if they passed the nominal significance in the joint model (P<0.05) (**Figure 1a**).

### Genome-wide PRS

The standard genome-wide AN PRS was also calculated for each individual in the PGC-ED cohorts using leave-one-out summary statistics of AN^19^. Clumping was performed on a genome-wide level using the PRSice default (--clump-kb 250, --clump-p 1, --clump-r2 0.1)^51^. The p-value thresholding for genome-wide PRS calculation was set at 1 (--bar-levels 1, --fastscore) to keep it the same threshold as for pathway PRS^50^.

### Pathway count in the top decile

Pathway PRS was first converted into deciles for all identified pathways for each individual (i.e., 1 as the lowest decile and 10 as the highest decile). A count of all pathways where the individual fell into the 10^th^ decile was created. For sensitivity analyses where the top percentage varied, we adjusted the counting accordingly. For example, to count Top_50%_ pathways, we used the number of pathways where the individual fell into either 6^th^,7^th^, 8^th^, 9^th^, or 10^th^ deciles.

### Correlation permutation

To assess whether the significant correlation between AN risk and number of pathways where the individual ranked at the top 10% was due to chance, we performed 10,000 permutations where we randomly selected 497 pathways (or 41 pathways) out of the 4,831 non-significant pathways (i.e., P>0.05 in the univariate model) to test their correlation and p-value with AN risk. The empirical p-value was calculated to assess if the correlation coefficient and associated p-value for our identified 497 pathways from the univariate model (or 41 pathways from the joint model) were significantly different from the null distribution (**Supplementary Fig. 8**).

### Pathway similarity by Jaccard Index

A Jaccard index (JI) is a measure of similarity across two sets^56^. In this study, we calculated JI for each pairwise pathway, which reflects the similarity between two pathways where the pathway would have a score of 0 if there is no gene overlap across the two pathways, and a score of 1 if all the genes in the two pathways are the same. In general, JI is the ratio of the intersection size (i.e., number of shared genes across the two pathways) over the union size (i.e., all genes in the two pathways)^56^, ranging from 0 to 1. Next, JI (i.e., similarity) across top pathways in both cases and controls were calculated and compared using the asymptotic two-sample Kolmogorov-Smirnov test, which is a nonparametric test to test whether the two groups had the same continuous distribution.

### Biological grouping of pathways

Within the 497 AN-associated pathways, we further classified these pathways into four different biological groups, which are brain-related, metabolic, immune/stress-related and reproduction-related. These biological groups were identified based on the name of the pathways themselves or their associated parent nodes given the tree structure of Gene Ontology^53^ using the GOfuncR package in R. The keywords used for identifying pathways for each biological group were (1) brain-related: “serotonin”, “dopamine”, “neuron”, “nervous”, “behavior”, “dendrite”, “neurogenesis”, “cognition”, “synapse”, “head”, “axon”, “synapse”, “synaptic”, “neurotransmitter”; (2) metabolic: “metabolic”; (3) immune/stress-related: “immune”, “defense”, “inflam”, “stress”; and (4) reproduction-related: “reproductive”, “reproduction”, “embryo”, respectively. A total of 49 brain-related pathways, 126 metabolic pathways, 39 immune/stress-related pathways, and 9 reproduction-related pathways were identified (**Supplementary Tables 12-15**). Manual review was also conducted for all 497 identified pathways to confirm that the biological grouping based on these keywords was sensible and thorough. Only two pathways were found through both immune and metabolic pathway searching, which are (1) DNA repair from GOBP; and (2) Regulation of retrograde protein transport (ER to cytosol) from GOBP. Sensitivity analysis showed that removing these two pathways did not substantially change the result (**Supplementary Fig. 16, Supplementary Table 17**). Within the 41 AN pathways identified from the joint model, a total of 4 brain-related pathways, 10 metabolic pathways, 1 immune/stress-related pathways, and 2 reproduction-related pathways were identified (Supplementary Table 5).

### Pairwise pathway correlation

Since most pathway PRS were normally distributed (e.g.,15.1% of 497 pathway PRS had P_skewness_<=0.05 [n = 75], 7.8% had P_kurtosis_<=0.05 [n=39]), Pearson correlation was used for pairwise correlation among all 497 pathways (**Supplementary Table 3**), all 41 pathways (**Figure 1b**) and pathways in the same and different biological groups (**Supplementary Fig. 7**). Sensitivity analysis was also performed using Spearman correlation, where we found a small difference in mean correlation across 497 pathway PRS (*r*_mean_difference_ = 0.007, P < 2.2×10^−16^). Given the small absolute difference and that the majority of pathway PRS were normally distributed, we kept the Pearson correlation as the primary correlation method.

### Pathway PRS residuals

To remove the possible confounding effect of AN PRS on pathway PRS, we computed residuals of pathway PRS adjusting for AN PRS, cohort, and six genetic PCs. We also computed deciles based on residuals of each pathway PRS for all individuals.

### Model performance measures

Nagelkerke R^2^, a pseudo-R^2^ measure for logistic regression, was calculated to examine the amount of variance in AN that was explained by pathway PRS and/or AN PRS. We first calculated Nagelkerke R^2^ of the full model with PRS and covariates as well as R^2^ of the null model with only covariates (i.e., cohort, 6 PCs). Next, the improvement in R^2^ explained by pathway PRS (or AN PRS) was calculated using the formula below.

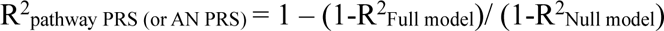

To put the improvement in R^2^ by the AN-associated pathway PRS into context, we also calculated R^2^ explained by a random set of 497 pathway PRS (or 41 pathway PRS), sampling from the 4,831 non-significant pathways over 100 permutations and computed empirical p-values for the significance of R^2^ based on our identified pathway PRS. Sensitivity test was also performed to examine whether gene set size or gene set database influenced the results by examining size-matched random pathways as well as random pathways from the REACTOME collection in the MSigDB database^54,57^ (version 7.4, Supplementary Results, **Supplementary Table 6**).

We also evaluated precision (i.e., positive predictive value) and recall (i.e., sensitivity) of using pathway PRS to predict AN cases and controls. Specifically, the (1) Precision-Recall (PR) curve was constructed along with (2) area under the curve (AUC, possible range from 0 to 1) and (3) AUC per SNP for AN PRS, 41 pathway PRS, and 497 pathway PRS, using the pracma package in R. A higher PR AUC indicates a better prediction performance. A higher PR AUC per SNP indicates a higher cost effectiveness for SNP genotyping. The number of SNPs for calculation of AN PRS, 41 pathway PRS, 497 pathway PRS was estimated using the largest cohort included in this study (i.e., the chop cohort, which recruited AN cases from US, Canada, UK, Germany and Italy and controls from the Children’s Hospital of Philadelphia, **Supplementary Table 1**)^43^.

Similarly, to evaluate sensitivity and specificity of pathway PRS, the Receiver Operating Characteristic (ROC) curve was plotted, using the pROC package in R. A higher ROC AUC indicates a better prediction performance, where ROC AUC = 1 indicates perfect prediction and ROC AUC = 0.5 indicates random guess.

### Comparison of PRSet and MAGMA

We also examined the performance of PRSet against MAGMA, a commonly used tool for gene set identification^35^. We first annotated SNPs to corresponding genes in MAGMA (v.1.0.8) using the same GTF file as in PRSet to define gene location. The same flanking region of 35 kb upstream and 10 kb downstream was used to include potential regulatory variants for each gene. Next, gene-based analysis was performed in the four selected cohorts with the largest effective sample sizes in our study (**Supplementary Table 1**), where effective sample size was calculated as 4/[(1/N_case_) + (1/N_control_)]. Gene-based analyses were calculated in two ways, (1) using the summary statistics of AN GWAS^19^ and (2) using individual genotype data (**Supplementary Table 1**). The 1000 Genome^58^ Phase 3 European genotype data was served as the reference panel for information on SNP linkage disequilibrium (LD). Next, built upon the output from the gene-based analysis, we performed pathway analysis using gene set information from the MSigDB database^54^ (version 7.4). Next, we calculated and compared replication rate across the four cohorts for significant gene sets identified by PRSet, MAGMA (based on raw genotype), and MAGMA (based on summary statistics). We defined replication rate as how many significant gene sets identified in one cohort were replicated in each of the other three cohorts (**Supplementary Fig. 18**). For the same set of MSigDB gene sets, we also assessed the performance of PRSet versus MAGMA (based on raw genotype) by examining the consistency of pathway effect size and p-values across cohorts (i.e., chop *versus* gns2, chop *versus* net1, chop *versus* usa1) using Pearson correlation (**Supplementary Figs. 19-21**).

### Role of gene significance in pathway significance

The gene-based p-values for 18,799 genes were retrieved from the supplementary table of the published AN GWAS which included 16,992 cases and 55,525 controls^19^. These genes covered 82% - 100% of the genes included in the 497 pathways. To assess whether there is a relationship between pathway significance and the most significant gene, we evaluated correlation between p-value of the most significant gene available and p-value of pathway PRS from both univariate models and the joint model (**Supplementary Fig. 3**).

### Statistical analysis

Significant difference in AN proportion in two different groups was assessed using a Two Proportions Calculator (see Web resources). To evaluate whether a model that included pathway PRS and AN PRS performed better than a model with only AN PRS, likelihood ratio test was performed to evaluate the model fit of these two generalized linear models. The pairwise differences in pathway PRS across cases and controls within each genetic risk group (i.e., low, middle, high genetic risk groups based on AN PRS), and across genetic risk groups within cases and controls were assessed using t-tests. To test whether within-function or across-function pathway pairs were associated with higher AN risk, AN case proportion differences across ten different groups were assessed. The ten groups include within-function pairs (i.e., brain-brain, metabolic-metabolic, immune-immune, reproduction-reproduction) and across-function pairs (i.e., brain-metabolic, brain-immune, brain-reproduction, metabolic-immune, metabolic- reproduction, immune-reproduction). The overall difference across ten groups was assessed by Kruskal-Wallis test (**Figure 4f, Supplementary Fig. 16**), followed by pairwise Wilcoxon rank sum tests across the ten groups adjusting for multiple testing correction via the Benjamini-Hochberg False Discovery Rate (FDR) method (**Supplementary Table 17**). The significance level for each correlation coefficient was computed based on linear regression, which generated the same p-value as the stat_cor() function in the ggpubr package in R.

### Reporting summary

Detailed information on the study design is available in the Nature Research Reporting Summary linked to this article.

## Data availability

Datasets from the Psychiatric Genomics Consortium Eating Disorders Working Group are available by requesting data access through the PGC data access portal (https://pgc.unc.edu/for-researchers/data-access-committee/data-access-portal/).

## Web resources

Collection of Gene Ontology Biological Process pathways in the Molecular Signatures Database, https://www.gsea-msigdb.org/gsea/msigdb/human/collections.jsp

PRSet tutorial, https://choishingwan.github.io/PRSice/prset_detail/ Comparing two independent population proportions,

https://stats.libretexts.org/Courses/Lake_Tahoe_Community_College/Introductory_Statistics_(OpenStax)_With_Multimedia_and_Interactivity/10%3A_Hypothesis_Testing_and_Confidence_Intervals_with_Two_Samples/10.04%3A_Comparing_Two_Independent_Population_Propor

## Code availability

The source code for the analyses is available and deposited in GitHub (https://github.com/xuj18/).

## Acknowledgements

This work was supported in part through the computational resources and staff expertise provided by the Dutch National supercomputer hosted at SURF, the Yale Center for Research Computing at Yale University, and the Scientific Computing at the Icahn School of Medicine at Mount Sinai. Research reported in this paper was supported by the Office of Research Infrastructure of the National Institutes of Health under award number S10OD018522 and S10OD026880. The content is solely the responsibility of the authors and does not necessarily represent the official views of the National Institutes of Health.

## Author contributions

L.M.H. and J.X. conceptualized and designed the study. J.X. and L.M.H. drafted the manuscript. J.X. and J.S.J. are the main analysts for this paper, with additional analytical help from C.L.. S.W.C. provided code support for PRSet. Members in the PGC-ED working group collected data in this study. P.F.O., J.G., and S.W.C. provided expertise in statistical method of pathway analysis. K.J.B. provided expertise in basic neuroscience. C.M.B. provided clinical expertise for eating disorders. All authors read and approved the final manuscript for submission.

## Supplementary information

The supplement includes additional results, seventeen supplementary tables and twenty-one supplementary figures.

