## Supplementary results and figures for "Polygenicity at the pathway level for anorexia nervosa"

### Supplementary Information

#### Table of Content


Supplementary Results ………………………………………………………………. 2
Supplementary Figures ……………………………………………………………… 5
Supplementary References ……………………………………………………….. 42


Supplementary Tables are included in a separate Excel spreadsheet.

#### Supplementary Results

##### A random set of 41 pathway PRS

The separation of mean pathway PRS by genome-wide AN PRS and AN case/control status was not observed by a random set of 41 pathway PRS out of non-significant pathways (4,831 pathways with $P > 0.05$ in the univariate models). The random set of 41 pathway PRS had little correlation with AN PRS (mean $r = 0.039$, SD = 0.026) and AN status (mean $r = 0.004$, SD = 0.007, **Supplementary Fig. 1**).

##### Permutation to examine $R^2$ explained by pathway PRS

To further examine whether the higher $R^2$ explained by pathway PRS was due to chance, we compared the model $R^2$ against 100 random permutations using non-significant gene sets from GOBP[1–3] and our manually curated psychiatric-related gene sets[4], and additionally examined whether significance in $R^2$ remains if we picked size-matched random pathways. Given the gene set size ranges from 7 to 1975 genes for 41 pathway PRS identified from the joint model, and that the gene set size for our non-significant pathways ranges from 1 to 537 genes, we assessed the $R^2$ of random pathways controlling for gene set size for a subset of pathways (n=15) that could be matched for size (+/- 10%). Specifically, we compared the $R^2$ of this subset of pathway PRS with (1) a random set of 15 pathways with matched size from the non-significant gene set list and (2) a random set of 15 pathways with matched size from the REACTOME collection in the Molecular Signatures Database (MSigDB version 7.4)[3,5] (**Supplementary Table 6**). Overall, we found that $R^2$ was significantly higher for our identified pathways than random pathways even after matching for gene set size regardless of which collection of random gene sets was used (GOBP + manual collection or REACTOME, **Supplementary Table 6**).

##### Assessment of prediction precision

We first derived the probability of each individual being a case based on the logistic regression model including AN PRS or pathway PRS, adjusting for cohort and 6 PCs. When we used the median probability as a threshold to classify individuals into cases and controls, the precision based on either AN PRS or pathway PRS was low, but the model based on 497 pathway PRS had relatively higher precision ($PPV_{497pathway}$ = 35.22%, $PPV_{41pathway}$ = 32.62%, $PPV_{AN\ PRS}$ = 32.43%, **Supplementary Table 8**). Next, we tested classification of people based on the 1st and 3rd quartiles of probability such that individuals below the 1st quartile were classified into controls, those above the 3rd quartile as cases, and those in between as uncertain. Using this threshold, the precision increased but remained low for all three PRS models, and the model with the relatively highest precision was still the 497 pathway PRS ($PPV_{497pathway}$ = 44.46%, $PPV_{41pathway}$ = 39.91%, $PPV_{AN\ PRS}$ = 40.44%, **Supplementary Table 9**). Both the precision-recall (PR) curve and the Receiver Operating Characteristic (ROC) curve, which plot precision, recall (i.e., sensitivity), and specificity across the whole range of probability threshold for a binary outcome (case/control), also indicated a greater prediction performance by the model with 497 pathway PRS, shown by the largest area under the curve (AUC) (**Figure 1d,e**).

#### Relationship between AN risk and pathway count in specific functional groups

In order to assess whether the non-significant correlation between reproduction-related pathways and AN risk ($r$ = 0.16, P=0.69) was due to low number of pathways in this group ($N_{reproduction}$ = 9), we downsized the number of pathways in each of the other groups to 9 to evaluate whether the significant relationship still persisted for brain, metabolic, and immune/stress-related pathways. We randomly sampled 9 pathways within each of the groups ($N_{brain}$ = 49, $N_{metabolic}$ = 126, $N_{immune/stress}$ = 39) and found that the significant relationship with AN risk remained ($r_{brain}$ = 0.94, $P_{brain}$ = 0.0014; $r_{metabolic}$ = 0.89, $P_{metabolic}$ = 0.019; $r_{immune/stress}$ = 0.85, $P_{immune/stress}$ = 0.0080), suggesting that the non-significant correlation between reproduction-related pathways and AN risk was not due to the technical effect of including a small number of pathways in this group.

#### Comparison of PRSet and MAGMA

We also assessed the performance of PRSet versus MAGMA by first examining their replication rate across cohorts. On average, PRSet has a higher average replication rate than MAGMA (PRSet: 8.5%, MAGMA using individual data: 5.1%, MAGMA using summary statistics: 5.1%, **Supplementary Fig. 18**). Next, we assessed whether the pathway effect size and p-value are more consistent across cohorts using PRSet versus MAGMA. We found that the pathway effect size was much more consistent across cohorts using PRSet (*r* ranges from 0.66 to 0.79) than MAGMA (*r* ranges from -0.02 to 0.01) (**Supplementary Fig. 19**). When we restricted the pathways to only those that passed nominal significance (P<0.05), the correlation of pathway effect size across cohorts improved for both MAGMA and PRSet, with PRSet having higher correlations across cohorts (PRSet: *r* ranges from 0.94 to 0.98, MAGMA using individual data: *r* ranges from 0.82 to 0.94, **Supplementary Fig. 20**). However, both PRSet and MAGMA had poor correlations for pathway p-value across cohorts (PRSet: *r* ranges from 0.013 to 0.102, MAGMA using individual data: *r* ranges from -0.004 to 0.015, **Supplementary Fig. 21**).

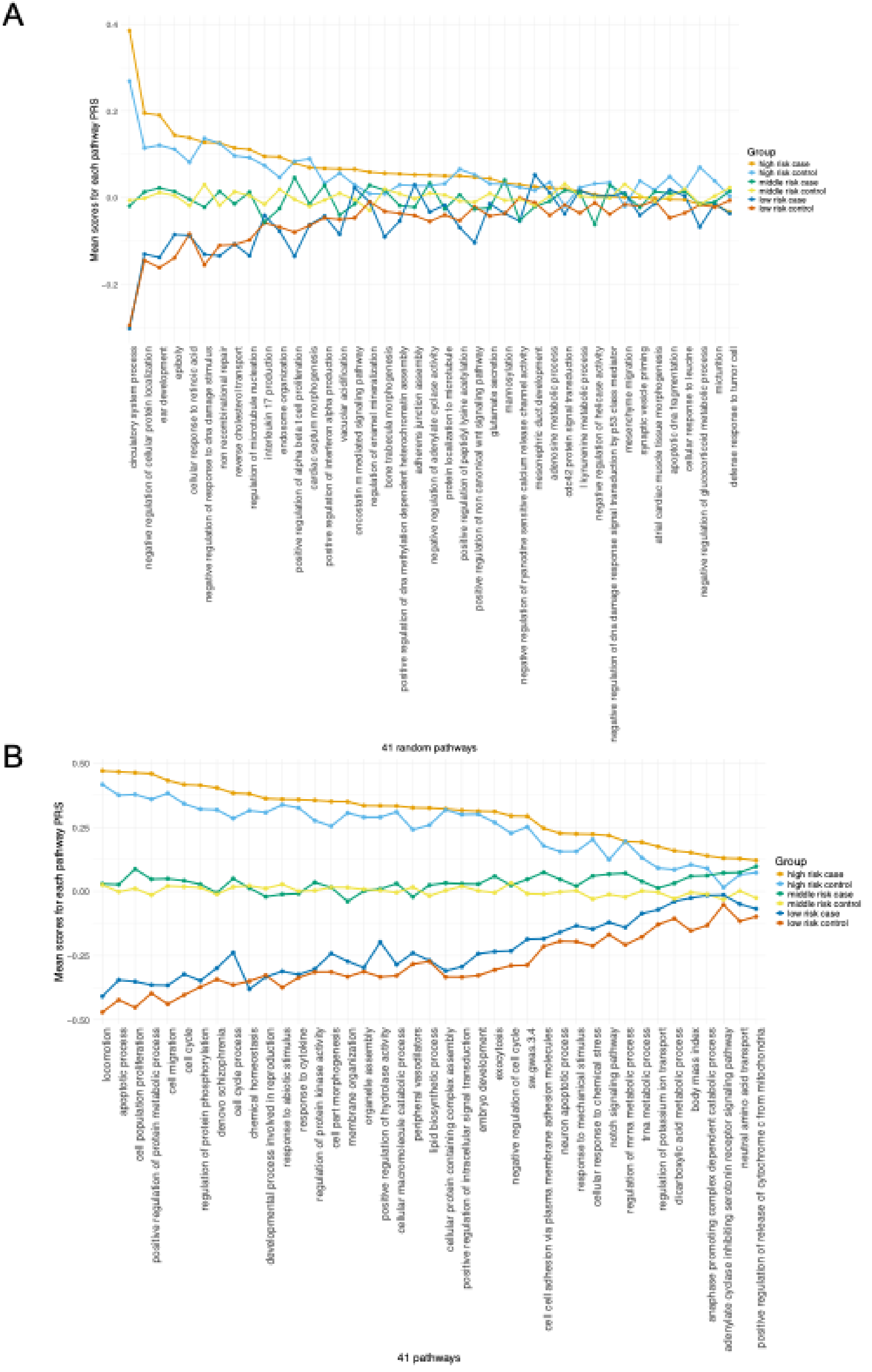
A
Mean scores for each pathway PRS
0.4
0.2
0.0
−0.2
Group
high risk case
high risk control
middle risk case
middle risk control
low risk case
low risk control
circulatory system process
negative regulation of cellular protein localization
ear development
epiboly
cellular response to retinoic acid
negative regulation of response to dna damage stimulus
non recombinational repair
reverse cholesterol transport
regulation of microtubule nucleation
interleukin 17 production
endosome organization
positive regulation of alpha beta t cell proliferation
cardiac septum morphogenesis
positive regulation of interferon alpha production
vacuolar acidification
oncostatin m mediated signaling pathway
regulation of enamel mineralization
bone trabecula morphogenesis
positive regulation of dna methylation dependent heterochromatin assembly
adherens junction assembly
negative regulation of adenylate cyclase activity
protein localization to microtubule
positive regulation of peptidyl lysine acetylation
positive regulation of non canonical wnt signaling pathway
glutamate secretion
mannosylation
negative regulation of ryanodine sensitive calcium release channel activity
mesonephric duct development
adenosine metabolic process
cdc42 protein signal transduction
l kynurenine metabolic process
negative regulation of helicase activity
negative regulation of dna damage response signal transduction by p53 class mediator
mesenchyme migration
synaptic vesicle priming
atrial cardiac muscle tissue morphogenesis
apoptotic dna fragmentation
cellular response to leucine
negative regulation of glucocorticoid metabolic process
micturition
defense response to tumor cell
41 random pathways
B
Mean scores for each pathway PRS
0.50
0.25
0.00
−0.25
−0.50
Group
high risk case
high risk control
middle risk case
middle risk control
low risk case
low risk control
locomotion
apoptotic process
cell population proliferation
positive regulation of protein metabolic process
cell migration
cell cycle
regulation of protein phosphorylation
denovo schizophrenia
cell cycle process
chemical homeostasis
developmental process involved in reproduction
response to abiotic stimulus
response to cytokine
regulation of protein kinase activity
cell part morphogenesis
membrane organization
organelle assembly
positive regulation of hydrolase activity
cellular macromolecule catabolic process
peripheral vasodilators
lipid biosynthetic process
cellular protein containing complex assembly
positive regulation of intracellular signal transduction
embryo development
exocytosis
negative regulation of cell cycle
sz.gwas.3.4
cell cell adhesion via plasma membrane adhesion molecules
neuron apoptotic process
response to mechanical stimulus
cellular response to chemical stress
notch signaling pathway
regulation of mrna metabolic process
trna metabolic process
regulation of potassium ion transport
dicarboxylic acid metabolic process
body mass index
anaphase promoting complex dependent catabolic process
adenylate cyclase inhibiting serotonin receptor signaling pathway
neutral amino acid transport
positive regulation of release of cytochrome c from mitochondria
41 pathways

**Supplementary Figure 1. A random set of 41 pathway PRS barely separates AN cases from controls.**
(A) The mean pathway PRS for each of 41 random pathway PRS is plotted, stratified by overall genetic risk (high, middle, low tertiles of AN PRS) and AN status (case/control).
(B) Same as **Figure 1c**, included here for visual comparison. The mean pathway PRS for each of 41 pathway PRS is plotted, stratified by overall genetic risk (high, middle, low tertiles of AN PRS) and AN status (case/control).
Abbreviations: AN=anorexia nervosa, PRS=polygenic risk score.

A

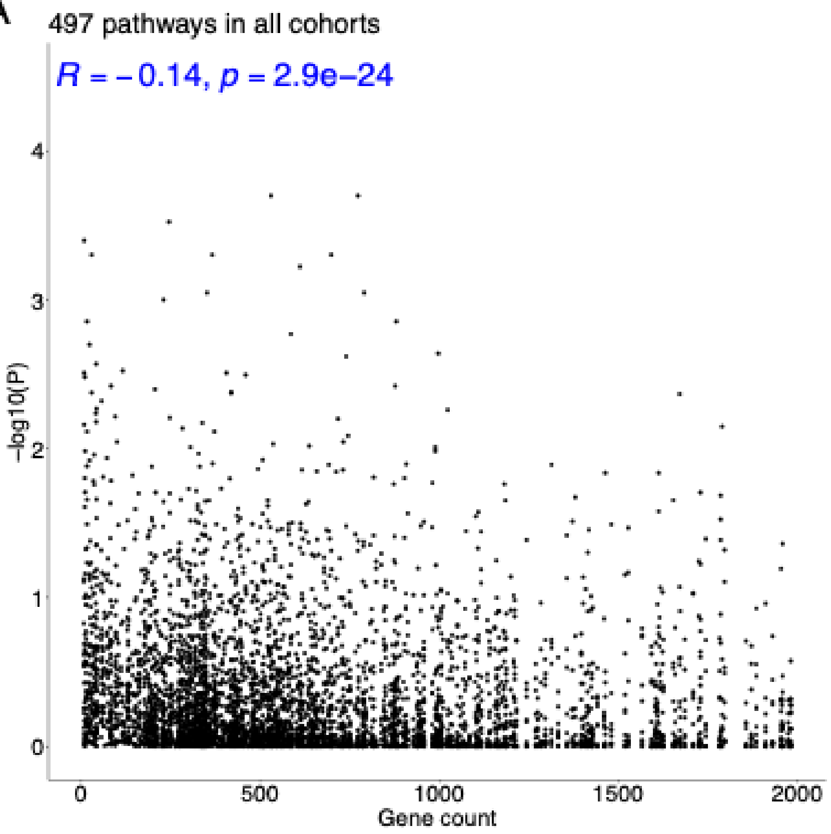

497 pathways in all cohorts
R = −0.14, p = 2.9e−24
−log10(P)
0
1
2
3
4
0
500
1000
1500
2000
Gene count


B

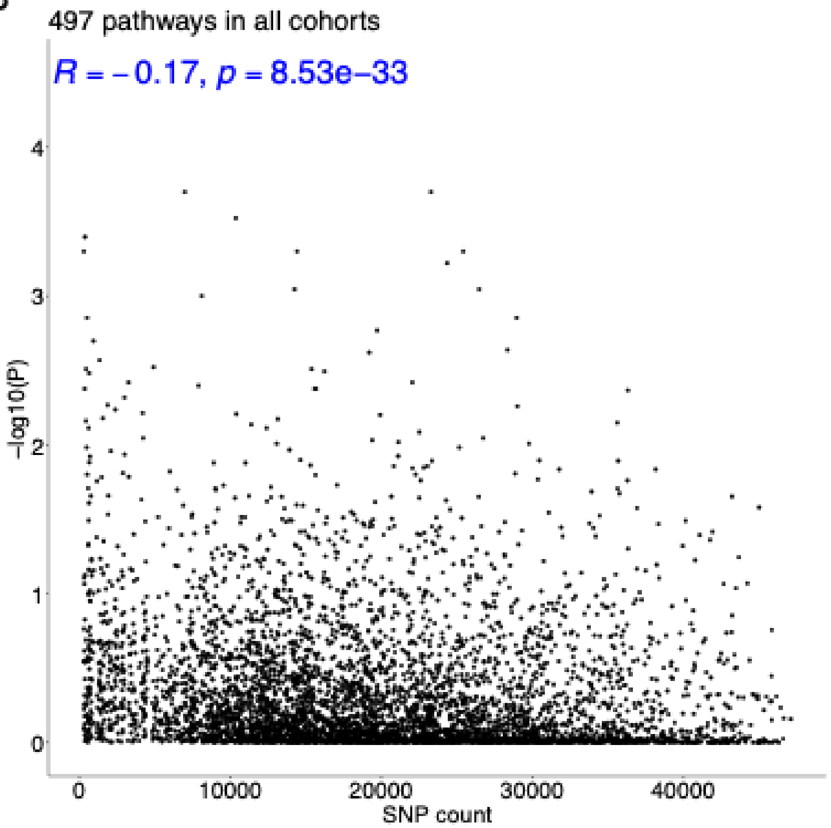

497 pathways in all cohorts
R = −0.17, p = 8.53e−33
−log10(P)
0
1
2
3
4
0
10000
20000
30000
40000
SNP count

**Supplementary Figure 2. Correlation between pathway size and pathway significance.**

(A) Correlation between pathway gene counts (x axis) and pathway significance (negative log10 p-value, y axis) for 497 significant pathways identified from the univariate models. The gene count of 497 pathway PRS ranges from 7 to 1982 genes. Pearson correlation and its associated p-value are shown in blue.

(B) Correlation between pathway SNP counts (x axis) and pathway significance (negative log10 p-value, y axis) for 497 significant pathways identified from the univariate models. The SNP count of 497 pathway PRS ranges from 293 to 47101 SNPs. Pearson correlation and its associated p-value are shown in blue.

Abbreviations: SNP=single nucleotide polymorphism.

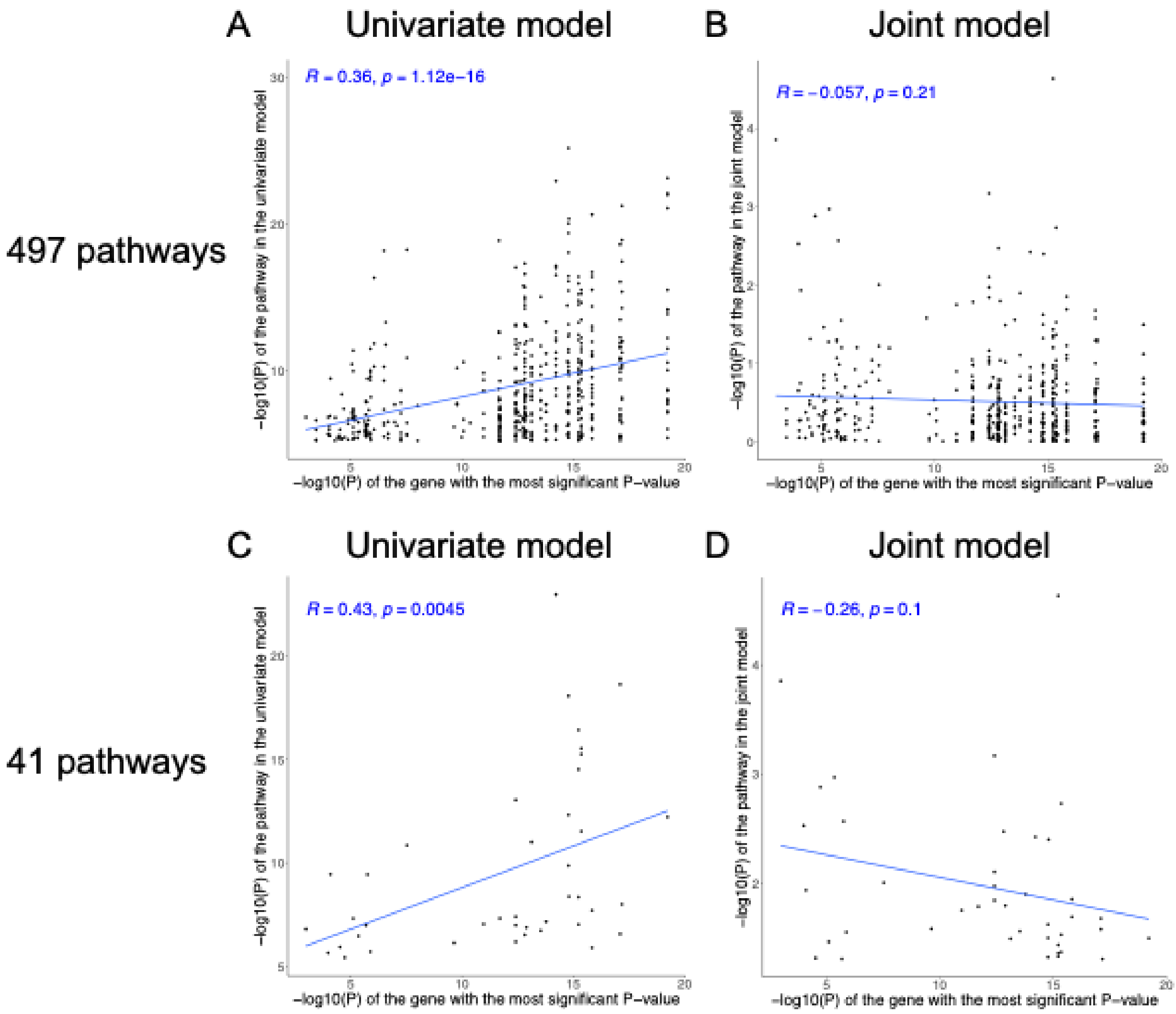


**Supplementary Figure 3. Correlation between pathway significance and the most significant gene within the pathway.**

(A) Correlation between the most significant gene (negative log10 p-value from Watson 2019[6], x axis) and pathway significance (negative log10 p-value, y axis) for 497 significant pathways identified from the univariate models. Pearson correlation, linear regression line, and the associated p-value are shown in blue. The p-value of each 497 pathway PRS from their corresponding univariate model was positively correlated with the p-value of the most significant gene within the pathway.

(B) Correlation between the most significant gene and pathway significance of 497 pathways from the joint model. The pathway p-value in the joint model was not correlated with the p-value of the most significant gene within the pathway.

(C) Correlation between the most significant gene and pathway significance for 41 significant pathways identified from the joint model. The p-value of each 41 pathway PRS from their corresponding univariate model was positively correlated with the p-value of the most significant gene within the pathway.

(D) Correlation between the most significant gene and pathway significance of 41 pathways from the joint model. The pathway p-value in the joint model was not correlated with the p-value of the most significant gene within the pathway. Abbreviations: PRS=polygenic risk score.

A

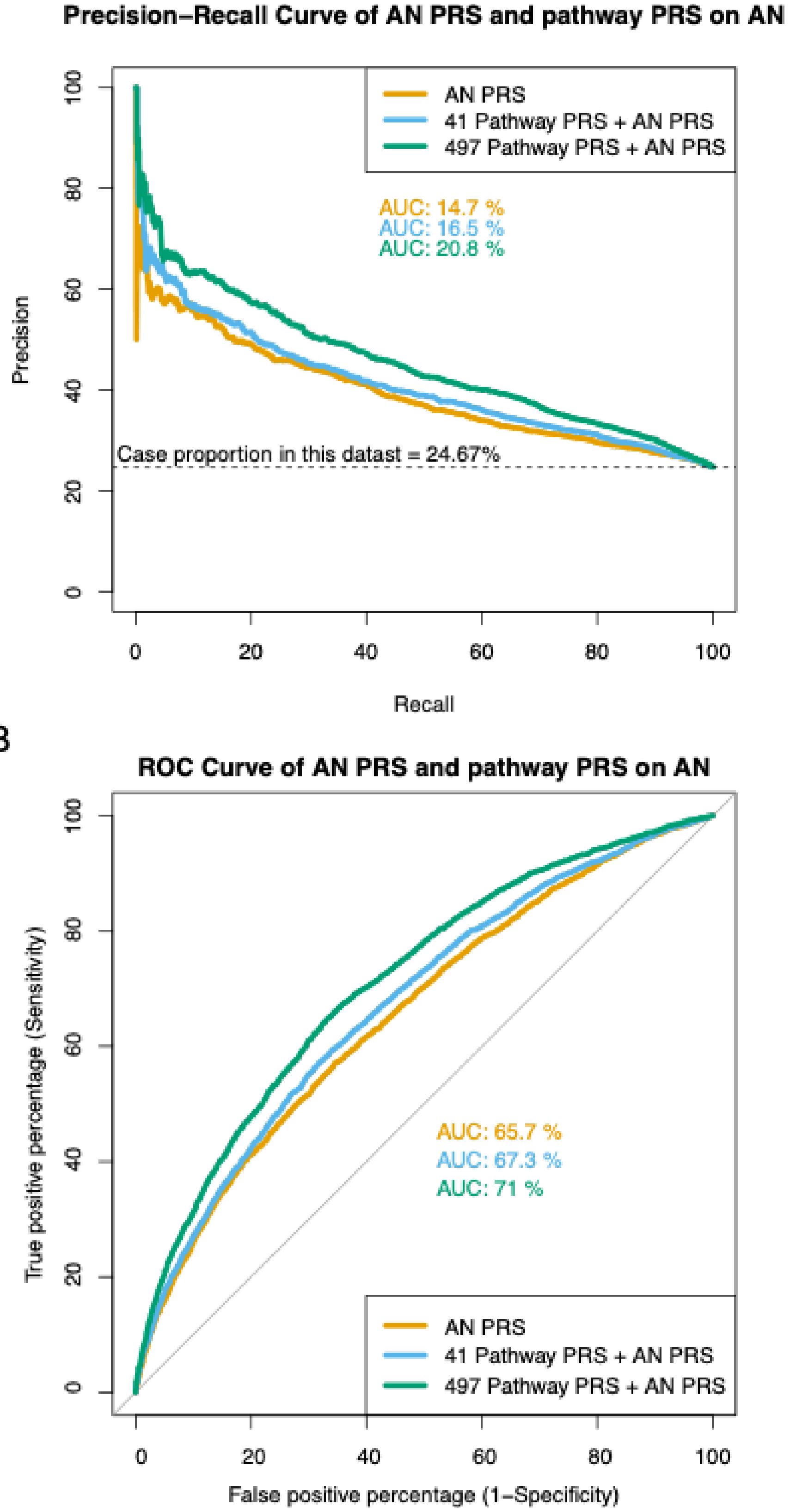

Precision−Recall Curve of AN PRS and pathway PRS on AN
AN PRS
41 Pathway PRS + AN PRS
497 Pathway PRS + AN PRS
AUC: 14.7 %
AUC: 16.5 %
AUC: 20.8 %
Case proportion in this datast = 24.67%
Precision
Recall
0
20
40
60
80
100
B
ROC Curve of AN PRS and pathway PRS on AN
AUC: 65.7 %
AUC: 67.3 %
AUC: 71 %
AN PRS
41 Pathway PRS + AN PRS
497 Pathway PRS + AN PRS
True positive percentage (Sensitivity)
False positive percentage (1−Specificity)

**Supplementary Figure 4. The highest area under the curve for both PR curve and ROC curve was achieved by 497 pathway PRS + AN PRS compared to 41 pathway PRS + AN PRS or AN PRS alone.**

(A) PR curve is plotted for AN PRS, 41 pathway PRS + AN PRS, and 497 pathway PRS + AN PRS, with AUC reported for each of them. The baseline line is y=24.67% as this equals the AN case proportion in the whole dataset. Here precision is defined as true positives over the sum of true positives and false positives, whereas recall (i.e., sensitivity) is defined as true positives over the sum of true positives and false negatives. A perfect prediction model would have a straight line at y=100% connected with x=100% at the top right corner.

(B) ROC curve is plotted for AN PRS, 41 pathway PRS + AN PRS, and 497 pathway PRS + AN PRS, with AUC reported for each of them. Here sensitivity (i.e., recall) is defined as true positives over the sum of true positives and false negatives, whereas specificity is defined as true negatives over the sum of true negatives and false positives. Therefore, 1 – specificity equals false positives over the sum of true negatives and false positives. A perfect prediction model would have a straight line at y=100% connected with x=0% at the top left corner. A random predictor with no discriminatory power would have an AUC of 50%.

Abbreviations: AN=anorexia nervosa, AUC=area under the curve, PR=precision-recall, PRS=polygenic risk score, ROC=receiver operating characteristic.

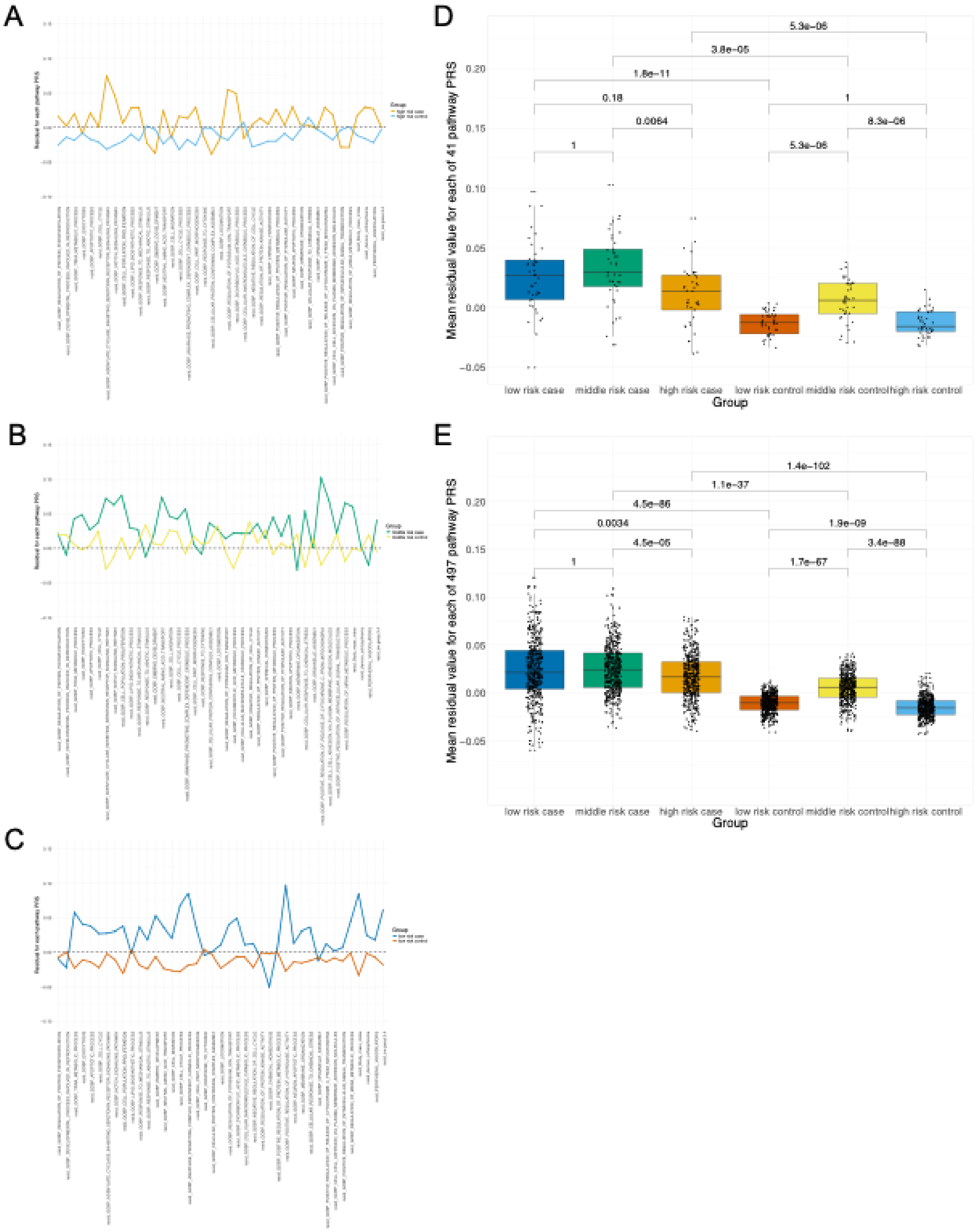
A
B
C
D
Mean residual value for each of 41 pathway PRS
5.3e−06
3.8e−05
1.8e−11
0.18
1
0.0064
8.3e−06
1
5.3e−06
low risk case
middle risk case
high risk case
low risk control
middle risk control
high risk control
Group
E
Mean residual value for each of 497 pathway PRS
1.4e−102
1.1e−37
4.5e−86
0.0034
1.9e−09
4.5e−05
3.4e−88
1
1.7e−67
low risk case
middle risk case
high risk case
low risk control
middle risk control
high risk control
Group

**Supplementary Figure 5. Pathway PRS residuals for AN cases and controls stratified by their genome-wide AN risk (high, middle, low).**
(A) Mean residuals of 41 pathway PRS in the high genetic risk group. Cases had mostly positive mean residuals, while controls had mostly negative mean residuals.
(B) Mean residuals of 41 pathway PRS in the middle genetic risk group. Cases had mostly positive mean residuals, while controls had mean residuals hovering around y=0.
(C) Mean residuals of 41 pathway PRS in the low genetic risk group. Cases had mostly positive mean residuals, while controls had mostly negative mean residuals.
(D) Boxplot that encompasses information of subfigure A-C, where each dot is the mean residual of each 41 pathway PRS across the six different groups by AN case/control status and overall genetic risk. Wilcoxon rank sum test was used to test for significant difference across each pairwise group with Bonferroni-corrected p-values shown for 9 group comparisons. Overall, high risk cases had smaller positive residuals than the other two case groups.
(E) Boxplot where each dot is the mean residual of each 497 pathway PRS across the six different groups by AN case/control status and overall genetic risk. Consistent with the finding observed for 41 pathway PRS, high risk cases had significantly smaller positive residuals than the other two case groups.
Abbreviations: AN=anorexia nervosa, PRS=polygenic risk score.

A

B

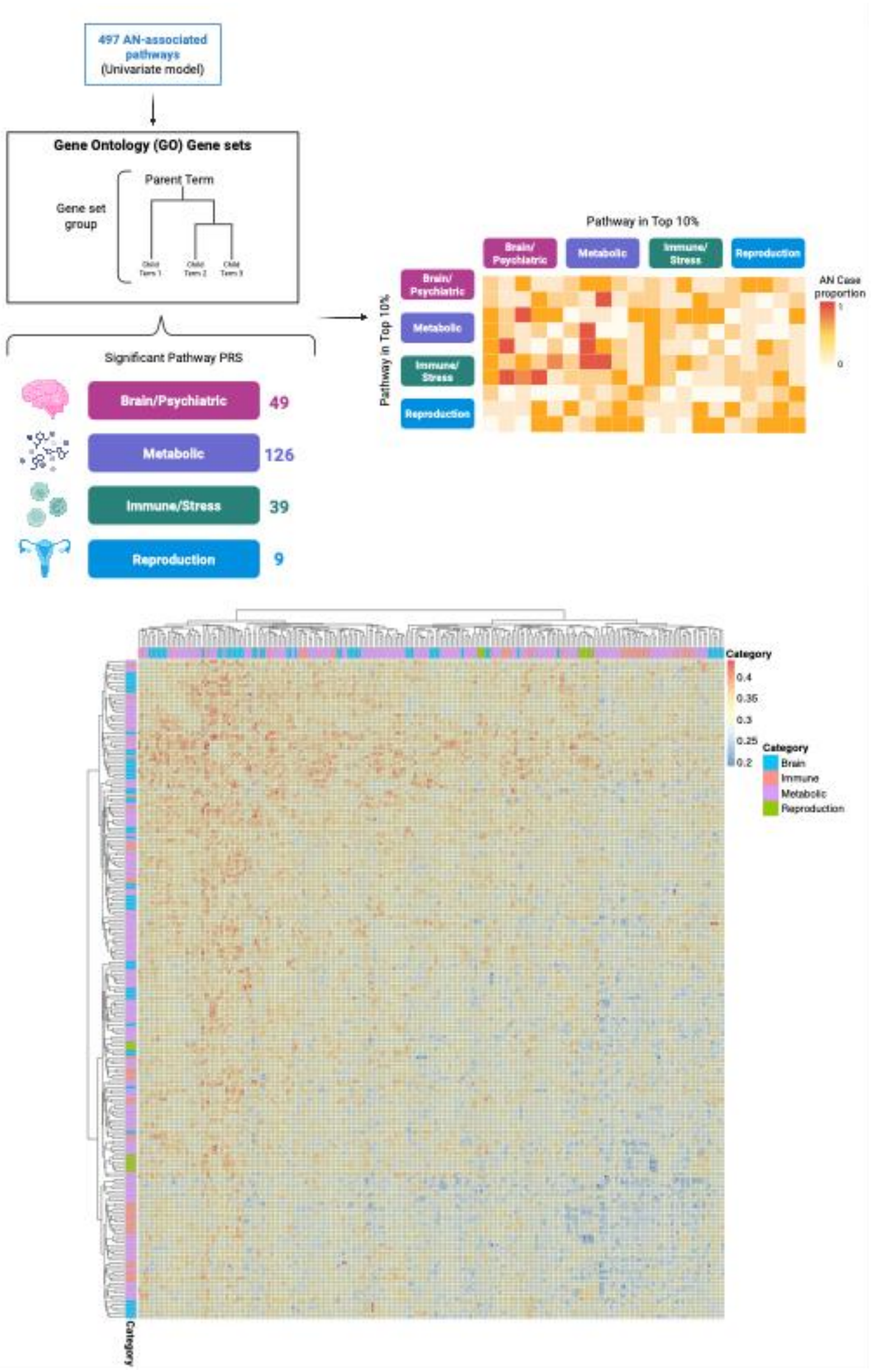

497 AN-associated pathways (Univariate model)
Gene Ontology (GO) Gene sets
Parent Term
Gene set group
Significant Pathway PRS
Brain/Psychiatric
49
Metabolic
126
Immune/Stress
39
Reproduction
9
Pathway in Top 10%
Brain/ Psychiatric
Metabolic
Immune/ Stress
Reproduction
Pathway in Top 10%
AN Case proportion
1
0
Category
0.4
0.35
0.3
0.25
0.2
Category
Brain
Immune
Metabolic
Reproduction
Category

**Supplementary Figure 6. Hierarchical clustering of individuals with top 10% genetic risks in two pathways in the four biological groups related to brain, metabolism, immune/stress and reproduction.**

(A) A toy figure to illustrate how the heatmap of AN case proportion for each pathway pair was generated. Four biological groups were identified based on the names of the 497 significant pathways themselves or their associated parent nodes given the tree structure of Gene Ontology using the GOfuncR package in R. Each cell in the heatmap contains individuals whose pathway PRS were ranked at top 10% for both pathways. AN case proportion was then calculated among these individuals in each cell. The heatmap shows clustered AN case proportion for pathway pairs across 49 brain-related pathways, 126 metabolic pathways, 39 immune/stress-related pathways, and 9 reproduction-related pathways. The exact AN case proportion in each cell is included in **Supplementary Table 16**. This figure was created using BioRender software (www.biorender.com).

(B) Hierarchical clustering heatmap of AN case proportion across pathway pairs. The proportion of anorexia nervosa ranges from 19.08% (bluer color) to 43.75% (redder color).

Abbreviations: AN=anorexia nervosa, PRS=polygenic risk score.

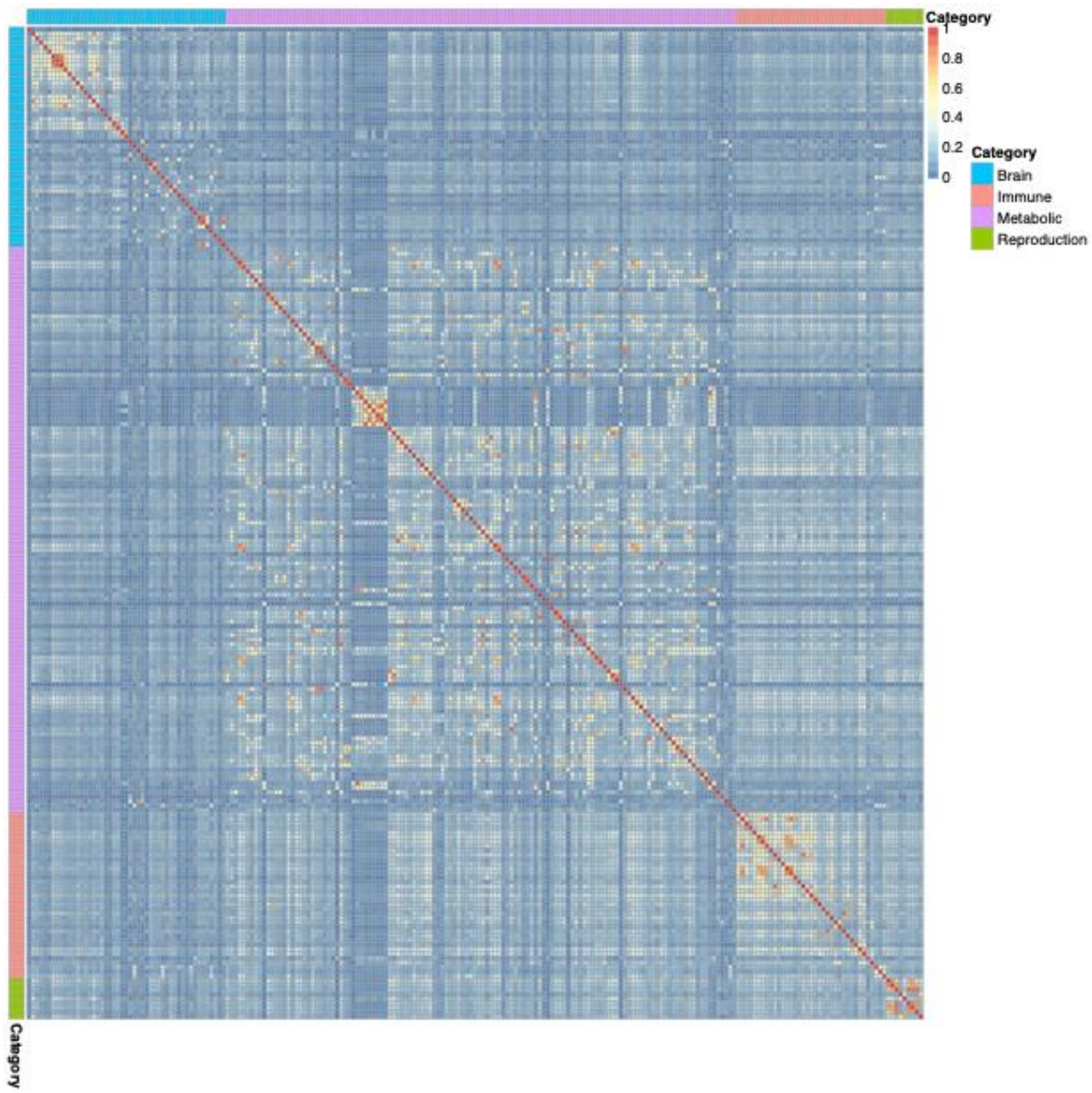


**Supplementary Figure 7. Correlation between two pathway PRS across the four biological groups related to brain, metabolism, immune/stress and reproduction**. In general, higher correlations are observed within each biological group (redder color) and lower correlations are observed across biological groups (bluer color). The mean Pearson correlation is 0.14 for the 221 pathway PRS in these four groups. Among them, only 0.8% are negative correlations with the most negative correlation of -0.02 for the pathway pair of pyrimidine nucleoside catabolic process (GOBP) and receptor internalization (GOBP). The highest correlation is 0.94 for the pathway pair of glycosyl compound metabolic process (GOBP) and nucleoside metabolic process (GOBP). Abbreviations: GOBP=Gene Ontology Biological Process, PRS=polygenic risk score.

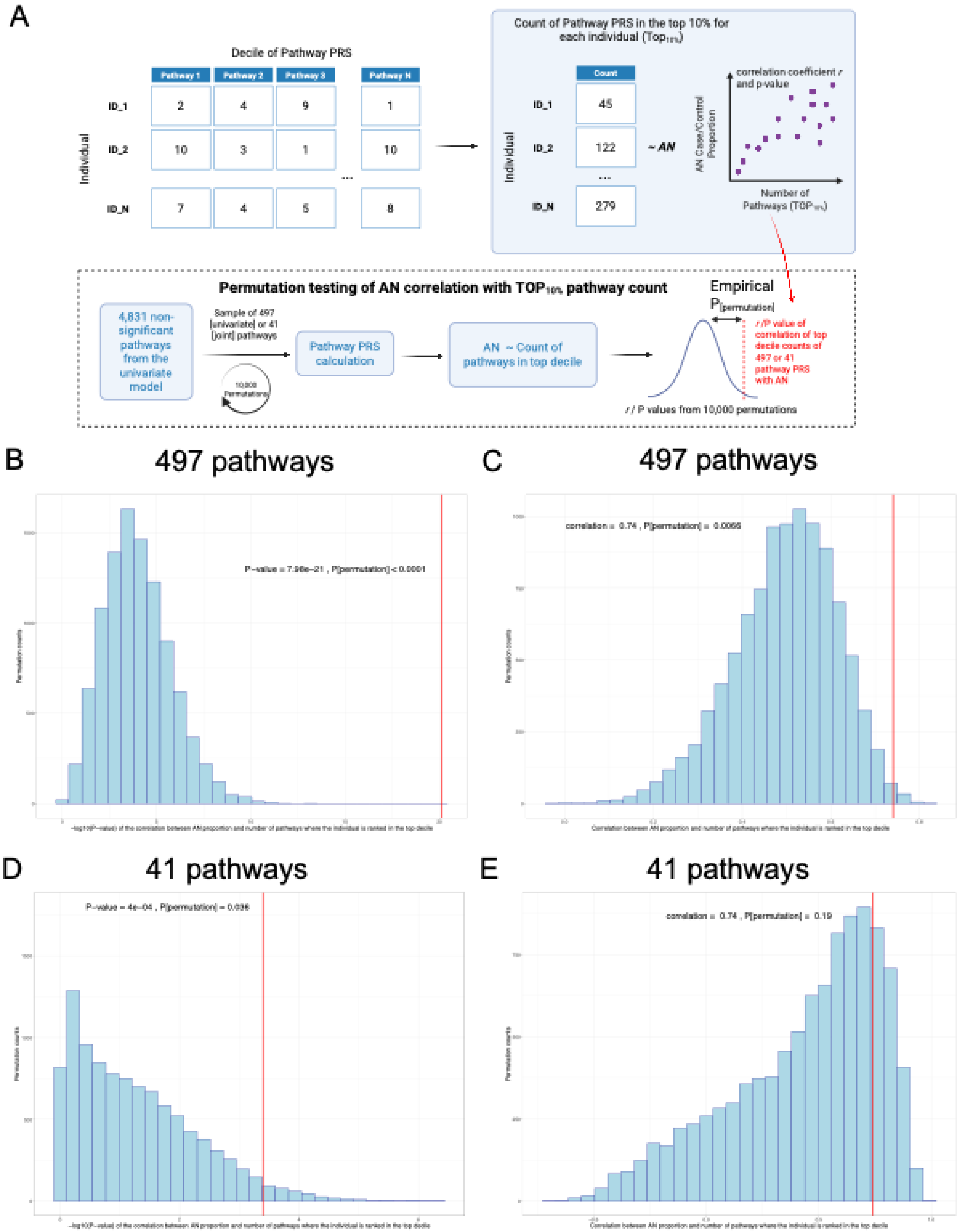

A
Decile of Pathway PRS
Pathway 1
Pathway 2
Pathway 3
Pathway N
Individual
ID_1
ID_2
ID_N
Count of Pathway PRS in the top 10% for each individual (Top10%)
Count
45
122
279
~ AN
correlation coefficient r and p-value
AN Case/Control Proportion
Number of Pathways (TOP10%)
Permutation testing of AN correlation with TOP10% pathway count
4,831 non-significant pathways from the univariate model
Sample of 497 (univariate) or 41 (joint) pathways
10,000 Permutations
Pathway PRS calculation
AN ~ Count of pathways in top decile
Empirical P[permutation]
r / P value of correlation of top decile counts of 497 or 41 pathway PRS with AN
r / P values from 10,000 permutations
B
497 pathways
P−value = 7.96e−21 , P[permutation] < 0.0001
C
497 pathways
correlation = 0.74 , P[permutation] = 0.0066
D
41 pathways
P−value = 4e−04 , P[permutation] = 0.036
E
41 pathways
correlation = 0.74 , P[permutation] = 0.19

**Supplementary Figure 8. The positive correlation between pathway counts in the top decile and AN case proportion was not due to chance.** The empirical p-values are significant for both the correlation coefficient (*r*) and p-value for 497 AN-associated pathways identified from the univariate models. While the empirical p-value for the correlation coefficient of 41 AN-associated pathways with AN proportion was not significant ($P_{permutation}$= 0.19), the corresponding correlation p-value was not due to chance ($P_{permutation}$= 0.036).
(A) A toy figure to illustrate how the permutation analysis was performed. First, significant pathway PRS identified in this study was converted into deciles for each individual. The number of pathways at the top decile for each individual was counted. Correlation was examined between AN case proportion and number of pathways at top 10% within 497 pathways and within 41 pathways. To test if this correlation is significantly different from the null distribution, we did 10,000 permutations where we randomly sampled 497 (or 41) pathway PRS from the 4,831 non-significant pathways 10,000 times to get a distribution of correlation coefficient (*r*) and correlation p-value between AN case proportion and top 10% pathway count when using randomly sampled pathway PRS. The empirical permutation p value was then calculated accordingly based on where the red dotted line falls in the distribution. This figure was created using BioRender software (www.biorender.com).
(B) Distribution of correlation p-values based on 10,000 permutations of a random set of 497 nonsignificant pathways, with the correlation p-value (red line, same value as **Figure 3b**) and its corresponding permutation p-value shown.
(C) Distribution of correlation coefficients based on 10,000 permutations of a random set of 497 nonsignificant pathways, with the correlation coefficient (red line, same value as **Figure 3b**) and its corresponding permutation p-value shown.
(D) Distribution of correlation p-values based on 10,000 permutations of a random set of 41 nonsignificant pathways, with the correlation p-value (red line, same value as **Figure 3c**) and its corresponding permutation p-value shown.
(E) Distribution of correlation coefficients based on 10,000 permutations of a random set of 41 nonsignificant pathways, with the correlation coefficient (red line, same value as **Figure 3c**) and its corresponding permutation p-value shown.
Abbreviations: AN=anorexia nervosa, PRS=polygenic risk score.

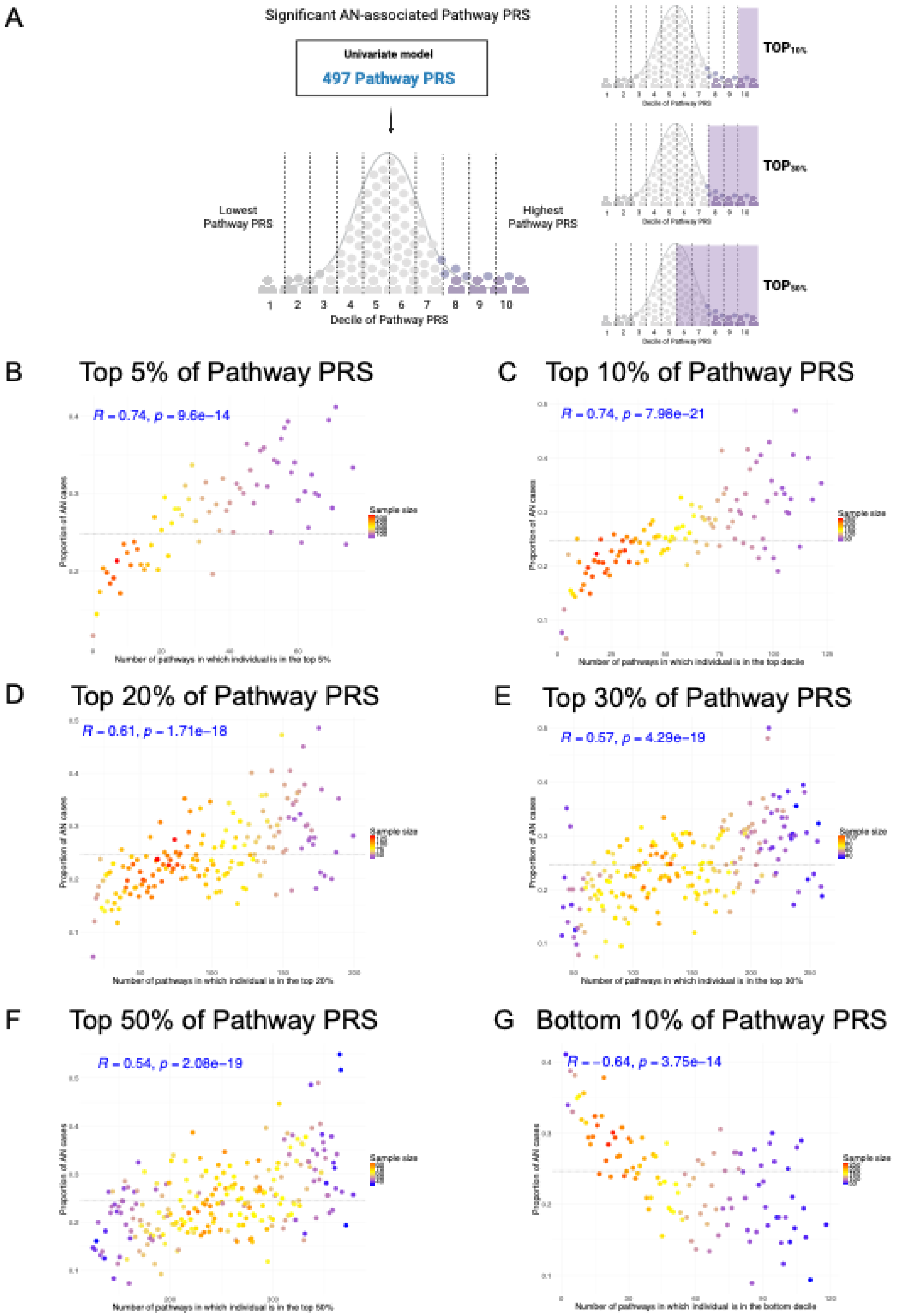
A
Significant AN-associated Pathway PRS
Univariate model
497 Pathway PRS
Lowest Pathway PRS
Highest Pathway PRS
1 2 3 4 5 6 7 8 9 10
Decile of Pathway PRS
TOP10%
TOP30%
TOP50%
B
Top 5% of Pathway PRS
R = 0.74, p = 9.6e−14
Proportion of AN cases
Number of pathways in which individual is in the top 5%
Sample size
C
Top 10% of Pathway PRS
R = 0.74, p = 7.98e−21
Proportion of AN cases
Number of pathways in which individual is in the top decile
Sample size
D
Top 20% of Pathway PRS
R = 0.61, p = 1.71e−18
Proportion of AN cases
Number of pathways in which individual is in the top 20%
Sample size
E
Top 30% of Pathway PRS
R = 0.57, p = 4.29e−19
Proportion of AN cases
Number of pathways in which individual is in the top 30%
Sample size
F
Top 50% of Pathway PRS
R = 0.54, p = 2.08e−19
Proportion of AN cases
Number of pathways in which individual is in the top 50%
Sample size
G
Bottom 10% of Pathway PRS
R = −0.64, p = 3.75e−14
Proportion of AN cases
Number of pathways in which individual is in the bottom decile
Sample size

**Supplementary Figure 9. Correlation of top/bottom pathway count with AN case proportion using different thresholds.** Each dot represents the group of individuals with certain number of pathways at the certain risk threshold. Only dots with more than 30 individuals are included for plotting. Pearson correlation and its associated p-value are shown at the top. The baseline AN proportion in the dataset is plotted as a dotted line (y=24.67%).
(A) A toy figure to illustrate how we defined pathway count at certain threshold. For example, a dot at x=50 in subfigure (C) includes all individuals who had 50 pathways that ranked at top 10% whereas a dot at x=50 in subfigure (E) includes all individuals who had 50 pathways that ranked at top 30%. And next, in each subfigure, AN case proportion was calculated for each dot. This figure was created using BioRender software (www.biorender.com).
(B) Relationship between number of pathways where the individual ranked at the top 5% within the 497 AN-associated pathways (x axis) and AN proportion (y axis) is plotted.
(C) Relationship between number of pathways where the individual ranked at the top 10% within the 497 AN-associated pathways (x axis) and AN proportion (y axis) is plotted (same as **Figure 3b** in the main manuscript).
(D) Relationship between number of pathways where the individual ranked at the top 20% within the 497 AN-associated pathways (x axis) and AN proportion (y axis) is plotted.
(E) Relationship between number of pathways where the individual ranked at the top 30% within the 497 AN-associated pathways (x axis) and AN proportion (y axis) is plotted.
(F) Relationship between number of pathways where the individual ranked at the top 50% within the 497 AN-associated pathways (x axis) and AN proportion (y axis) is plotted.
(G) Relationship between number of pathways where the individual ranked at the bottom 10% within the 497 AN-associated pathways (x axis) and AN proportion (y axis) is plotted.
Abbreviations: AN=anorexia nervosa.

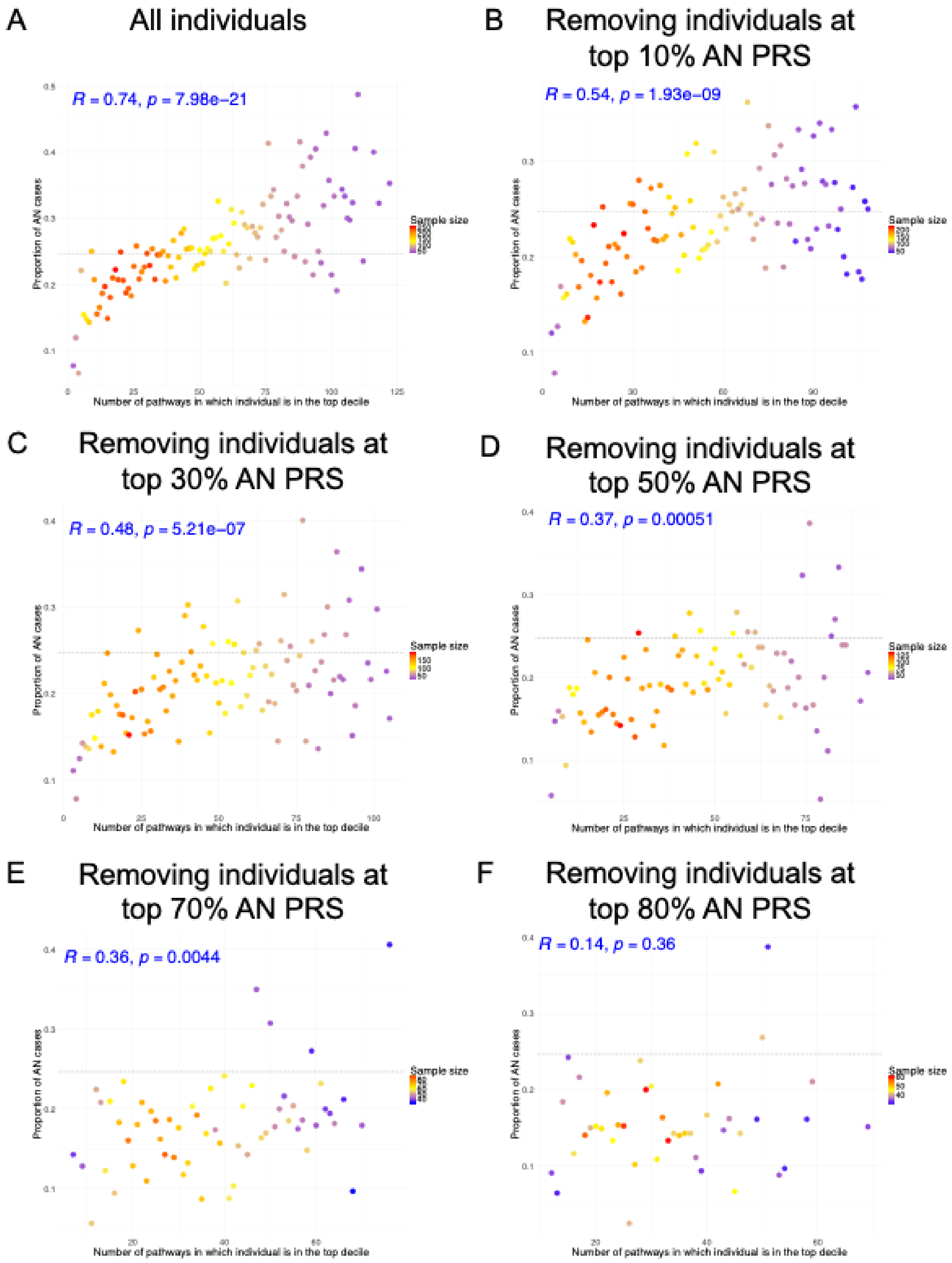
A
All individuals
R = 0.74, p = 7.98e−21
Proportion of AN cases
Number of pathways in which individual is in the top decile
Sample size
B
Removing individuals at top 10% AN PRS
R = 0.54, p = 1.93e−09
Proportion of AN cases
Number of pathways in which individual is in the top decile
Sample size
C
Removing individuals at top 30% AN PRS
R = 0.48, p = 5.21e−07
Proportion of AN cases
Number of pathways in which individual is in the top decile
Sample size
D
Removing individuals at top 50% AN PRS
R = 0.37, p = 0.00051
Proportion of AN cases
Number of pathways in which individual is in the top decile
Sample size
E
Removing individuals at top 70% AN PRS
R = 0.36, p = 0.0044
Proportion of AN cases
Number of pathways in which individual is in the top decile
Sample size
F
Removing individuals at top 80% AN PRS
R = 0.14, p = 0.36
Proportion of AN cases
Number of pathways in which individual is in the top decile
Sample size

**Supplementary Figure 10. Correlation of pathway count at top 10% with AN case proportion after removing people with top AN PRS defined by different thresholds.** Each dot represents the group of individuals with certain number of pathways at top 10% risk. Only dots with more than 30 individuals are included for plotting. Pearson correlation and its associated p-value are shown at the top. The baseline AN proportion in the whole dataset is plotted as a dotted line (y=24.67%).
(A) Relationship between pathway count where the individual ranked at the top 10% within the 497 AN-associated pathways (x axis) and AN proportion (y axis) is plotted for all individuals (same as **Figure 3b** in the main manuscript).
(B) The same relationship is plotted after removing individuals with top 10% AN PRS.
(C) The same relationship is plotted after removing individuals with top 30% AN PRS.
(D) The same relationship is plotted after removing individuals with top 50% AN PRS.
(E) The same relationship is plotted after removing individuals with top 70% AN PRS.
(F) The same relationship is plotted after removing individuals with top 80% AN PRS.
Abbreviations: AN=anorexia nervosa, PRS=polygenic risk score.

A Random pathways, removing individuals at top 70% AN PRS

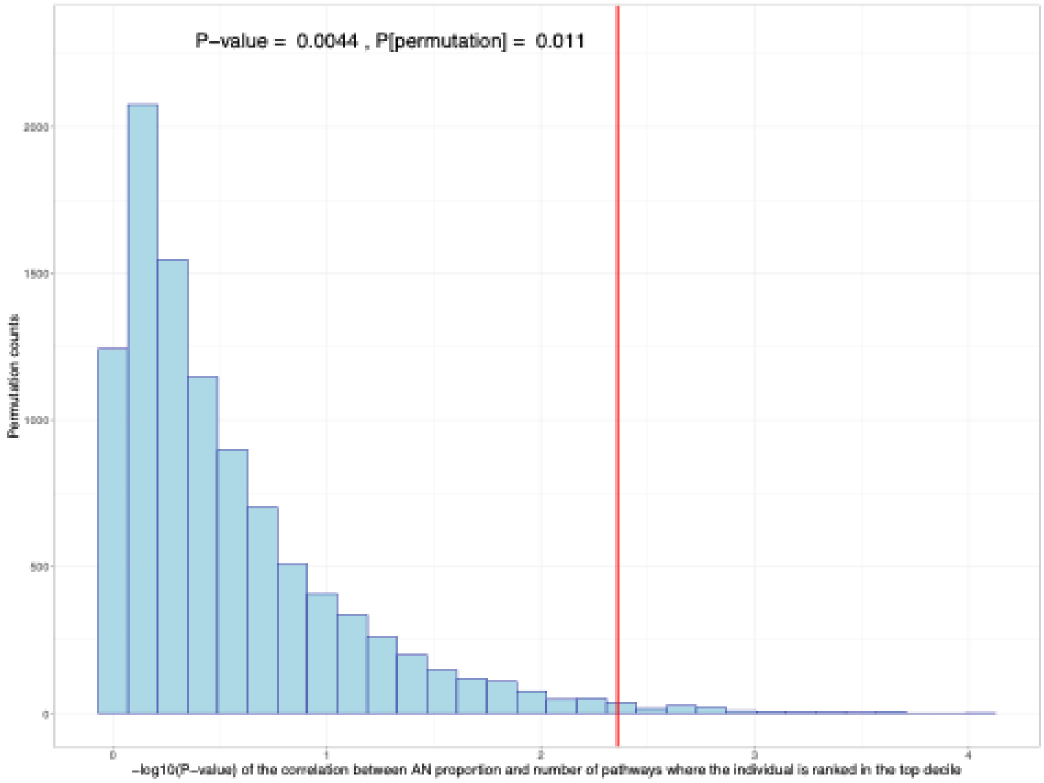


B Random pathways, removing individuals at top 70% AN PRS

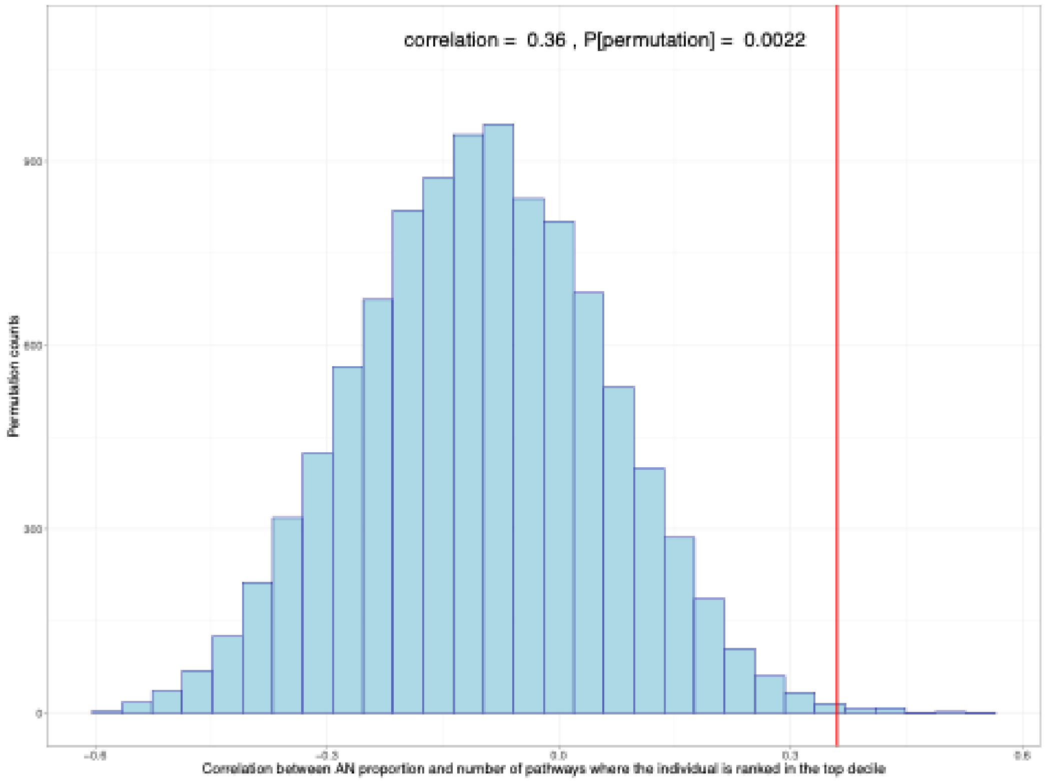

**Supplementary Figure 11. The significantly positive correlation between pathway counts in the top decile and AN case proportion among individuals with low genome-wide genetic risk (i.e., bottom 30% AN PRS) was not due to chance.** The empirical p-values are significant for both the correlation coefficient and p-value for 497 AN-associated pathways.
(A) Distribution of correlation p-values based on 10,000 permutations of a random set of 497 nonsignificant pathways, with the correlation p-value (red line, same value as **Supplement Figure 10e**) and its corresponding permutation p-value shown.
(B) Distribution of correlation coefficients based on 10,000 permutations of a random set of 497 nonsignificant pathways, with the correlation coefficient (red line, same value as **Supplement Figure 10e**) and its corresponding permutation p-value shown.
Abbreviations: AN=anorexia nervosa, PRS=polygenic risk score.

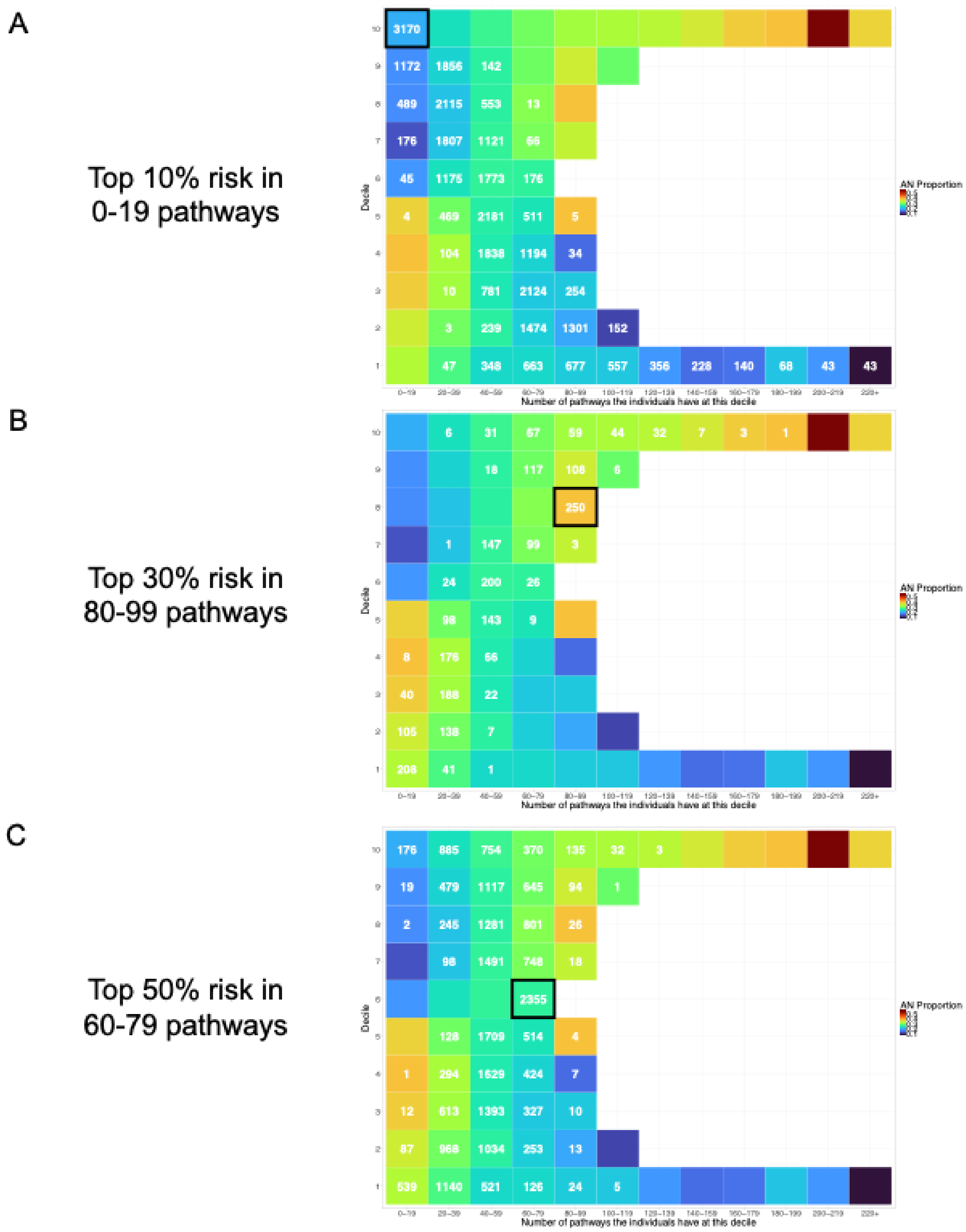
A
Top 10% risk in
0-19 pathways
B
Top 30% risk in
80-99 pathways
C
Top 50% risk in
60-79 pathways
Decile
Number of pathways the individuals have at this decile
AN Proportion

**Supplementary Figure 12. Three example scenarios to illustrate the distribution of individuals in one specific tile at other pathway risk levels.** The higher AN proportion in individuals with above-average risks in many pathways than those with concentrated risk in a few top 10% pathways could be because these individuals also had more pathway risks at top 10% in the meanwhile.

(A) For 3170 individuals with top 10% risk for 0-19 pathways, all these individuals had bottom 10% pathway risk for at least 20 pathways, and 43 of them (1.4%) had bottom 10% genetic risk for 220 pathways or more. At each decile, each row adds up to 3170, except that there was 1 individual at the 6$^{th}$ decile for 80-99 pathways, 1 individual at the 3$^{rd}$ decile for 100-119 pathways, and 1 individual at the 2$^{nd}$ decile for 120-139 pathways that are not shown as tiles with a total count less than 10 are not plotted.

(B) For 250 individuals with top 20-30% risk for 80-99 pathways, all these individuals also had top 10% pathway risk for at least 20 pathways, and 87 of them (34.8%) had top 10% genetic risk for 100 pathways or more. At each decile, each row adds up to 250, except that there was 1 individual at the 9$^{th}$ decile for 120-139 pathways that is not shown as tiles with a total count less than 10 are not plotted.

(C) For 2355 individuals with top 40-50% risk for 60-79 pathways,176 of them (7.5%) overlapped with individuals at top 10% risk for 0-19 pathways, which means that 92.5% of them also had top 10% pathway risk for at least 20 pathways, and 35 of them (1.5%) had top 10% genetic risk for 100 pathways or more. At each decile, each row adds up to 2355.

Abbreviations: AN=anorexia nervosa.

A

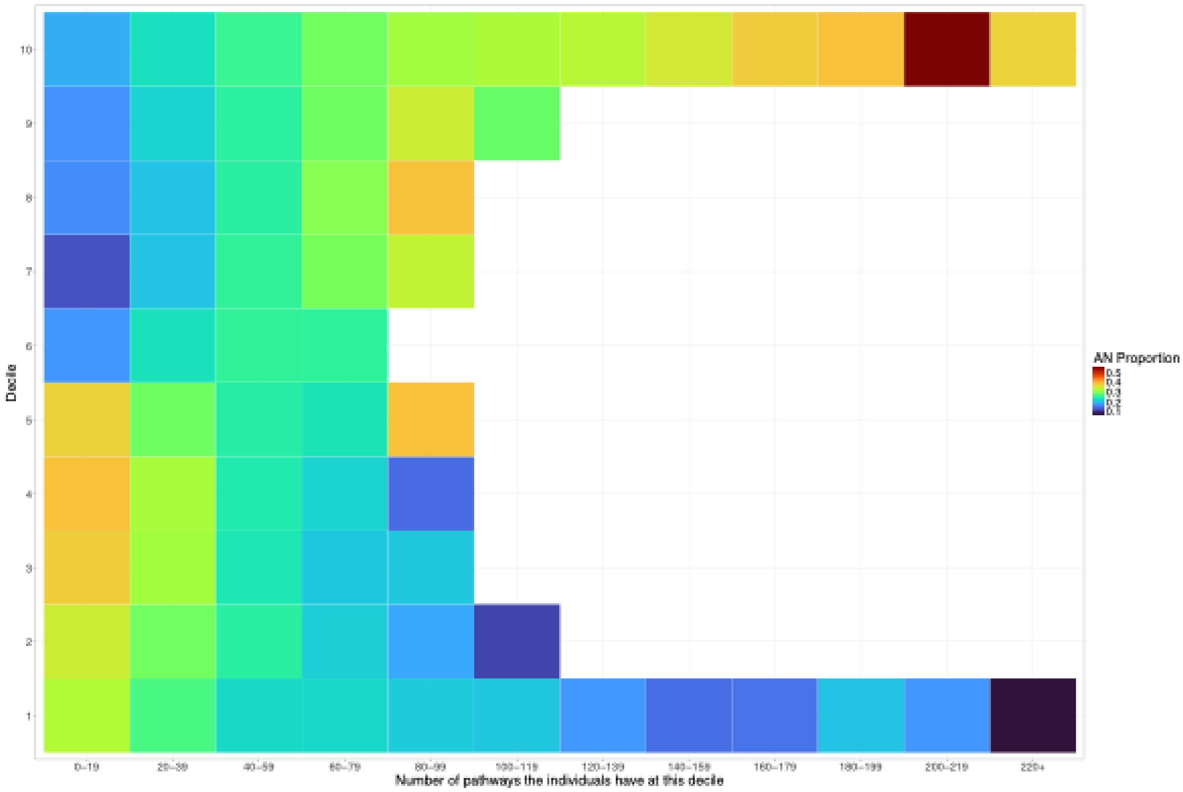

Decile
10
9
8
7
6
5
4
3
2
1
0–19
20–39
40–59
60–79
80–99
100–119
120–139
140–159
160–179
180–199
200–219
220+
Number of pathways the individuals have at this decile
AN Proportion
0.5
0.4
0.3
0.2
0.1


B

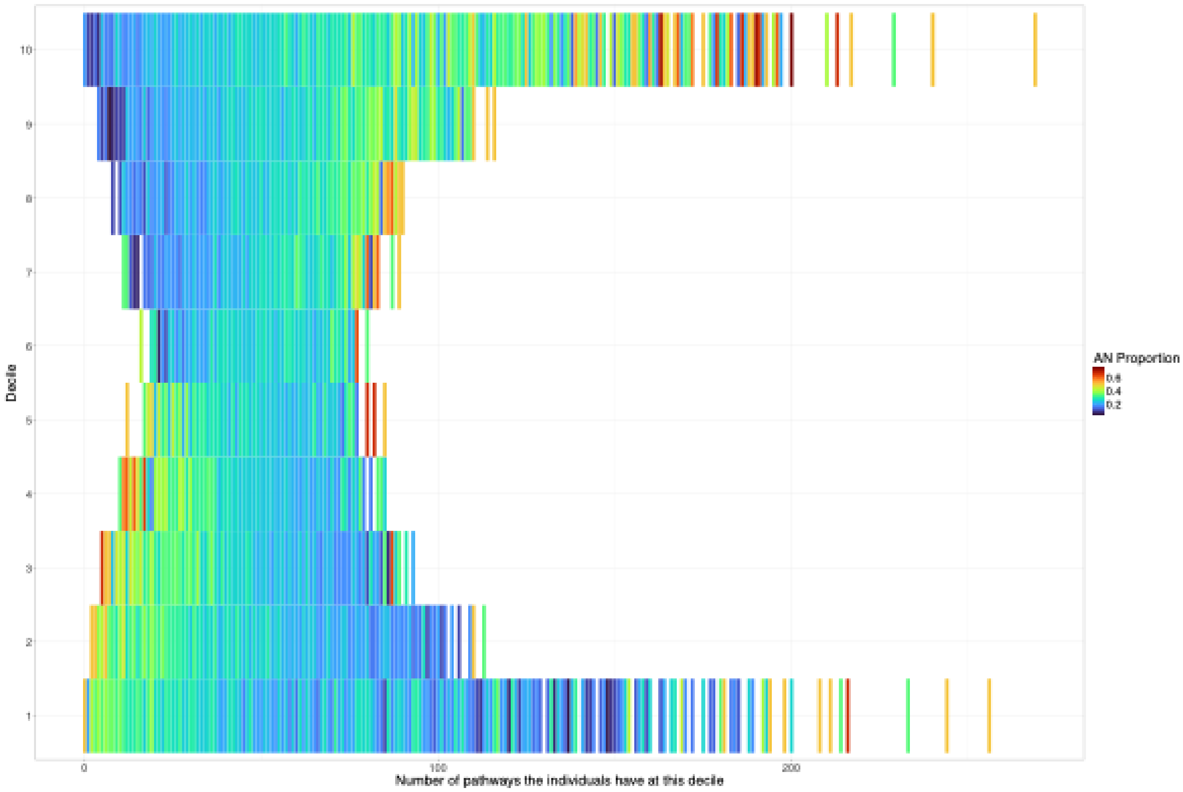

Decile
10
9
8
7
6
5
4
3
2
1
0
100
200
Number of pathways the individuals have at this decile
AN Proportion
0.6
0.4
0.2

**Supplementary Figure 13. AN proportion at different pathway risk levels across the range of pathway counts.**
**(A)** Same as **Figure 3g** in the main manuscript. Tiles with a total head count less than 10 are not plotted.
**(B)** Higher resolution by plotting the pathway count (x axis) as a continuous variable. The range of sample size in each tile ranges from 2 to 684 individuals, with a mean sample size of 142.9.
Abbreviations: AN=anorexia nervosa.

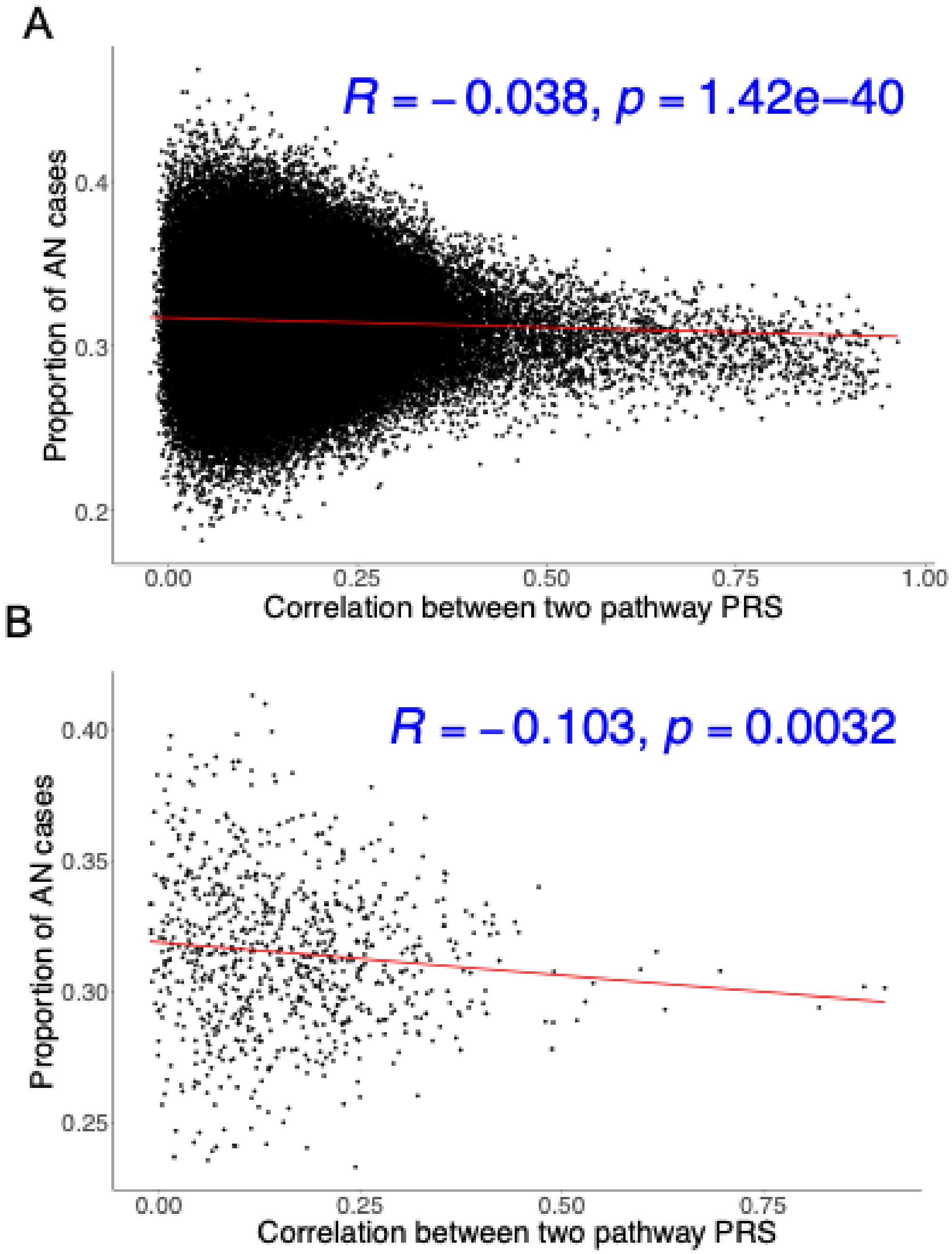


**Supplementary Figure 14. Relationship between pathway correlation and AN risk.**
(A) Pairwise similarity between pathways based on correlation (x axis) and AN case proportion (y axis) using 497 pathways PRS. Pearson correlation and its associated p-value are shown in blue.
(B) Pairwise similarity between pathways based on correlation (x axis) and AN case proportion (y axis) using 41 pathways PRS.
Abbreviations: AN=anorexia nervosa, PRS=polygenic risk score.

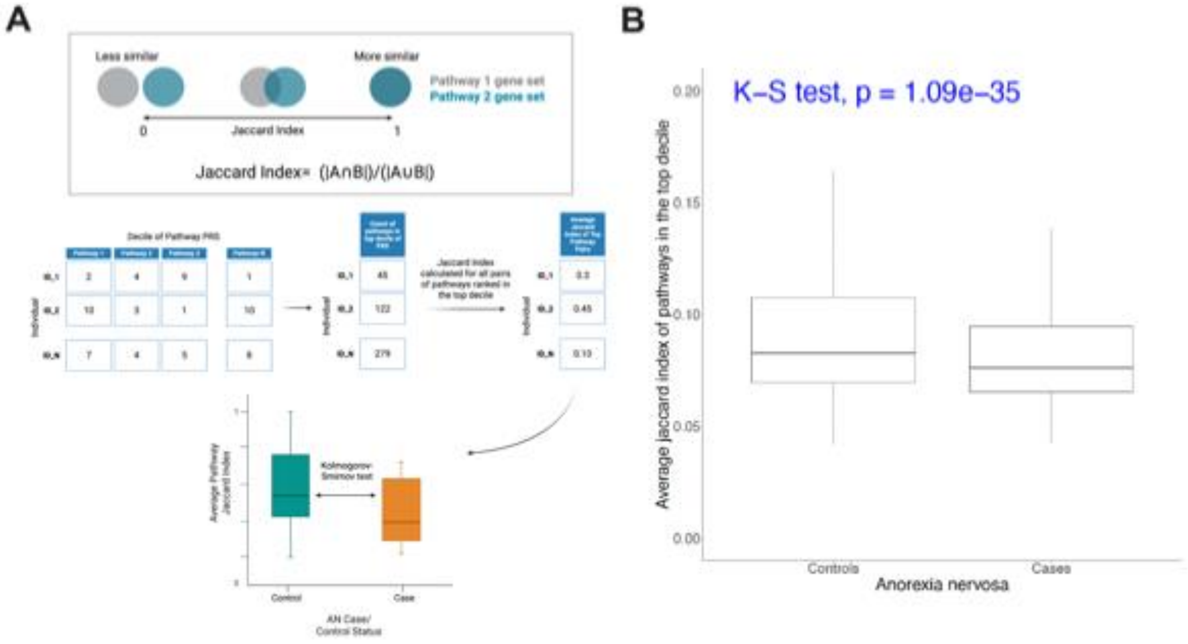


**Supplementary Figure 15. Relationship between pathway similarity and AN risk.**
(A) A toy figure to illustrate how the Jaccard index (JI) was calculated between two pathways, with JI=0 indicating no overlap and JI=1 indicating complete overlap. Thus, a lower JI indicates lower similarity (i.e., greater diversity). We first calculated how many pathways at top 10% each person had. And then among these top pathways, we calculated JI for each pair of pathways for each individual. Next, an average JI was calculated for each individual. We then plotted the average JI distribution in AN cases and controls and examined whether there was a significant difference between cases and controls in terms of top pathway similarity using the K-S test. This figure was created using BioRender software (www.biorender.com).
(B) Similarity of top pathways in cases versus controls based on JI within the 497 pathways.
Abbreviations: AN=anorexia nervosa, JI=Jaccard Index, K-S test=Kolmogorov-Smirnov test, PRS=polygenic risk score.

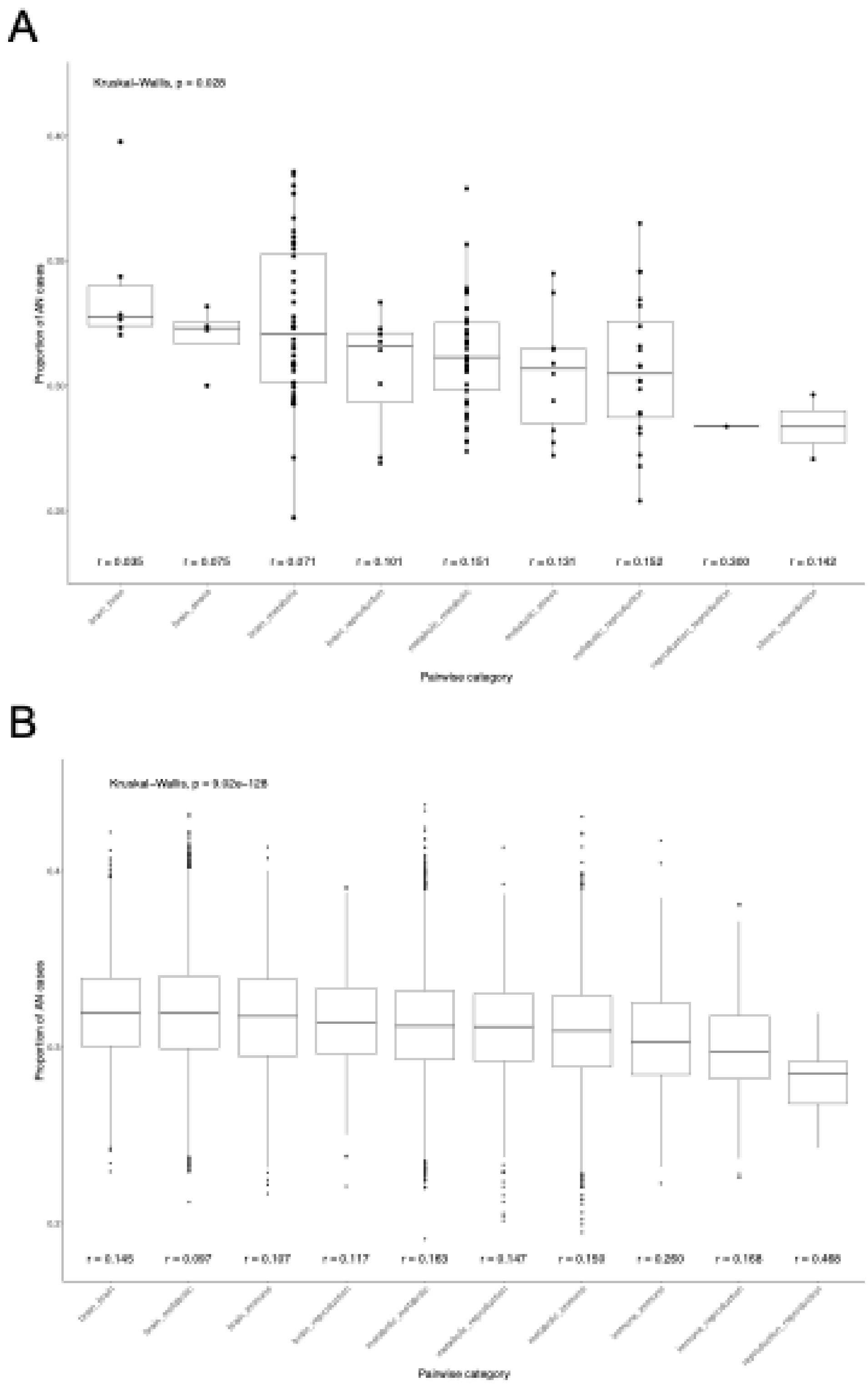
A
B
Proportion of AN cases
Pairwise category

**Supplementary Figure 16. Relationship of pathway function with AN risk using a smaller subset of pathways.**
(A) Boxplot of AN proportion distribution for pairwise pathways within and across functions for 17 pathways in the 4 biological groups among 41 AN-associated pathways from the joint model. The immune/stress-related pathway only has one relevant pathway included (i.e., cellular response to chemical stress in GOBP) and therefore this category is named only as stress (x axis) in this plot. The order is based on median AN proportion per category. Significance of overall difference across all groups was examined by Kruskal-Wallis test. P-values for all pairwise comparisons across 9 categories are included in **Supplemental Table 17**. The mean correlation for each group is shown at the bottom.
(B) Boxplot of AN proportion distribution for pairwise pathways within and across functions for 221 pathways in the 4 biological groups among 497 AN-associated pathways from the univariate models. Two overlapping pathways were removed from the metabolic and immune/stress-related groups. The two pathways are DNA Repair (GOBP) and Regulation of Retrograde Protein Transport, ER to Cytosol (GOBP), which were classified as both metabolic and immune/stress-related pathways. The ranking of pathway categories here is consistent with that in **Figure 4f**.
Abbreviations: AN=anorexia nervosa, GOBP=Gene Ontology Biological Process; PRS=polygenic risk score.

A PRSet (35 kb / 10kb)

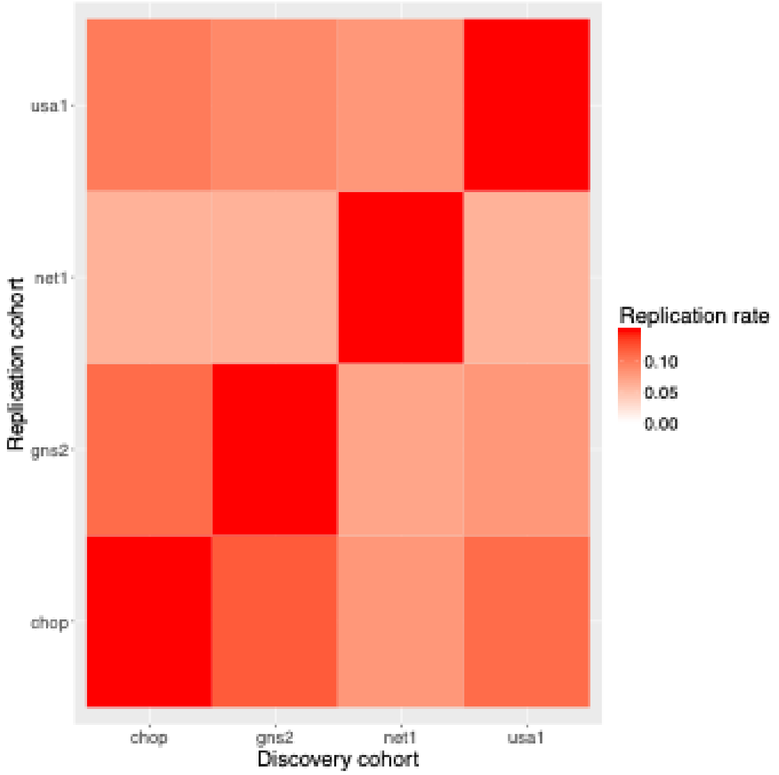


B PRSet (1 mb / 1 mb)

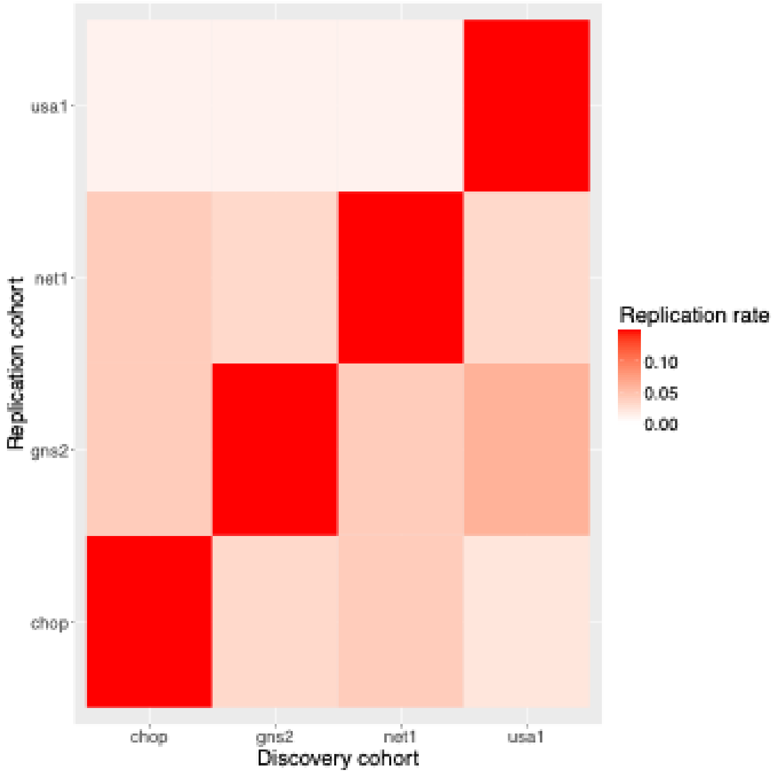

**Supplementary Figure 17. PRSet has a higher replication rate when using 35 kb / 10 kb gene flanking region.** The replication rate was calculated for each discovery-replication cohort pair (e.g., chop as the discovery cohort, gns2 as the replication cohort).
(A) Replication rate when 35kb upstream and 10kb downstream were used as gene flanking region. Replication rate ranges from 6% - 12% across cohorts.
(B) Replication rate when 1mb upstream and 1mb downstream were used as gene flanking region. Replication rate ranges from 1% - 6% across cohorts.
Abbreviations: kb=kilobase, mb=megabase.

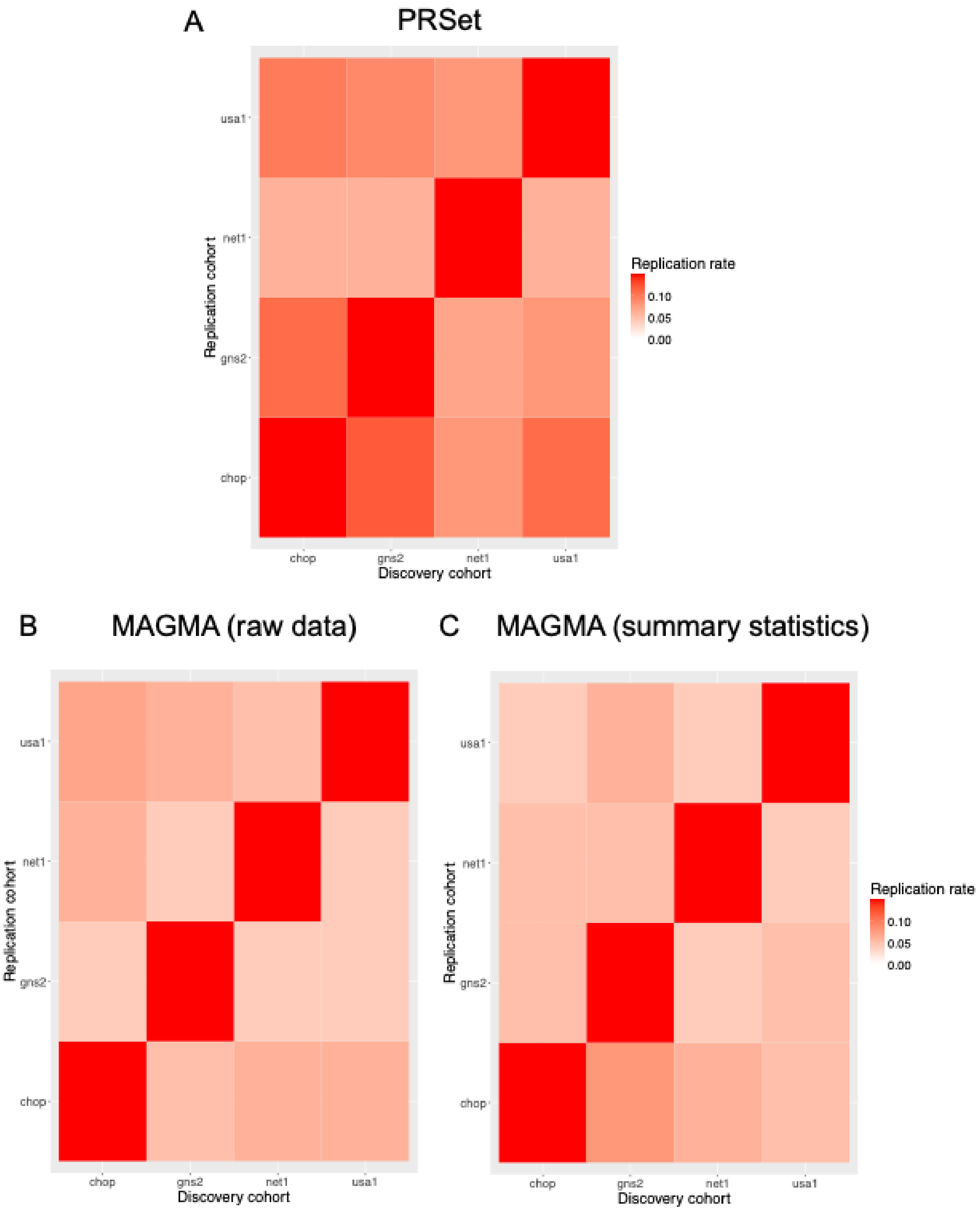

A
PRSet
B
MAGMA (raw data)
C
MAGMA (summary statistics)
Replication cohort
Discovery cohort
usa1
net1
gns2
chop
Replication rate
0.10
0.05
0.00

**Supplementary Figure 18. PRSet has a higher replication rate across cohorts than MAGMA[7].** The gene flanking region was set as 35 kilobases upstream and 10 kilobases downstream for both PRSet and MAGMA. The replication rate was calculated for each discovery-replication cohort pair (e.g., chop as the discovery cohort, gns2 as the replication cohort).
(A) Replication rate ranges from 6% - 12% using PRSet.
(B) Replication rate ranges from 4% - 8% using MAGMA with individual genotype data.
(C) Replication rate ranges from 4% - 7% using MAGMA with GWAS summary statistics.
Abbreviation: GWAS=genome-wide association study.

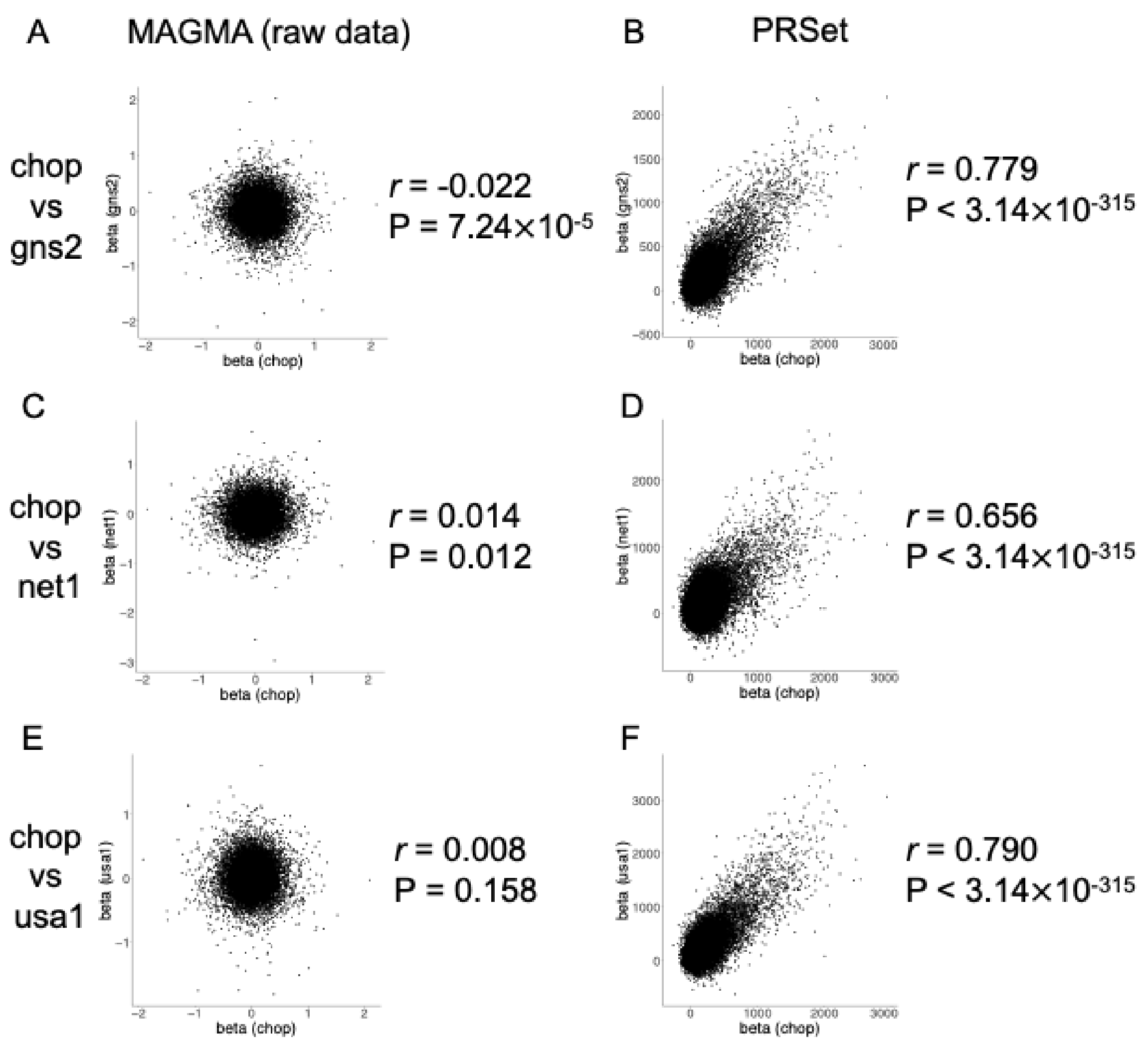


**Supplementary Figure 19. PRSet has a higher correlation of gene set effect size across discovery-replication cohort pairs compared to MAGMA.** Individual genetic data in these cohorts were used for gene set analysis by PRSet and MAGMA. Pearson correlation and its associated p-value are shown for each plot.
(A) Correlation between beta coefficient of the association between pathway and AN status in chop and beta coefficient in gns2 using MAGMA.
(B) Correlation between beta coefficient of the association between pathway and AN status in chop and beta coefficient in gns2 using PRSet.
(C) Correlation between beta coefficient of the association between pathway and AN status in chop and beta coefficient in net1 using MAGMA.
(D) Correlation between beta coefficient of the association between pathway and AN status in chop and beta coefficient in net1 using PRSet.
(E) Correlation between beta coefficient of the association between pathway and AN status in chop and beta coefficient in usa1 using MAGMA.
(F) Correlation between beta coefficient of the association between pathway and AN status in chop and beta coefficient in usa1 using PRSet.
Abbreviation: AN=anorexia nervosa.

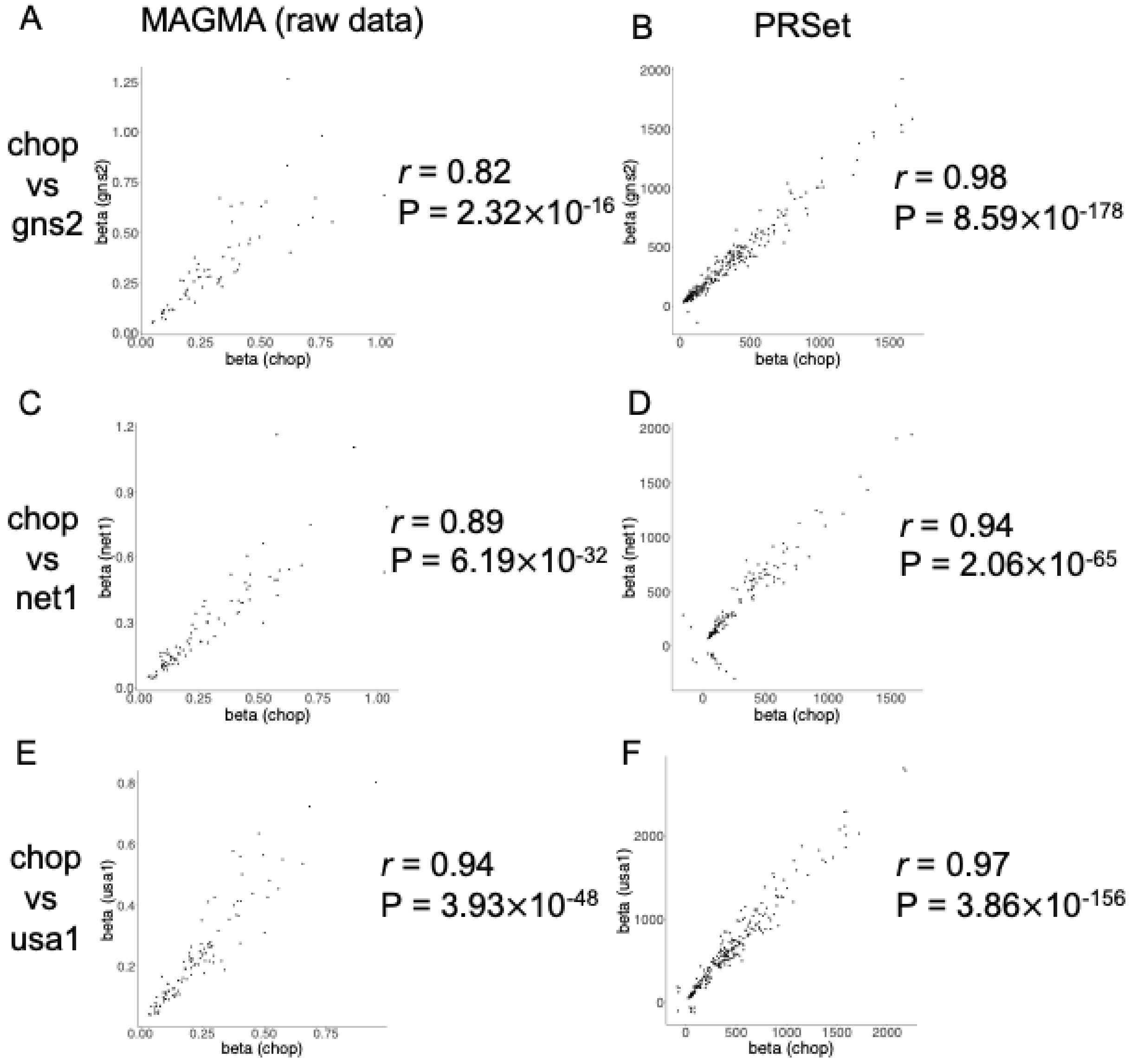


**Supplementary Figure 20. PRSet has a higher correlation of gene set effect size across discovery-replication cohort pairs compared to MAGMA when restricting to nominally significant gene sets ($P < 0.05$).** Individual genetic data in these cohorts were used for gene set analysis by PRSet and MAGMA. Pearson correlation and its associated p-value are shown for each plot.

(A) Correlation between beta coefficient of the association between pathway and AN status in chop and beta coefficient in gns2 using MAGMA.

(B) Correlation between beta coefficient of the association between pathway and AN status in chop and beta coefficient in gns2 using PRSet.

(C) Correlation between beta coefficient of the association between pathway and AN status in chop and beta coefficient in net1 using MAGMA.

(D) Correlation between beta coefficient of the association between pathway and AN status in chop and beta coefficient in net1 using PRSet.

(E) Correlation between beta coefficient of the association between pathway and AN status in chop and beta coefficient in usa1 using MAGMA.
(F) Correlation between beta coefficient of the association between pathway and AN status in chop and beta coefficient in usa1 using PRSet.
Abbreviation: AN=anorexia nervosa.

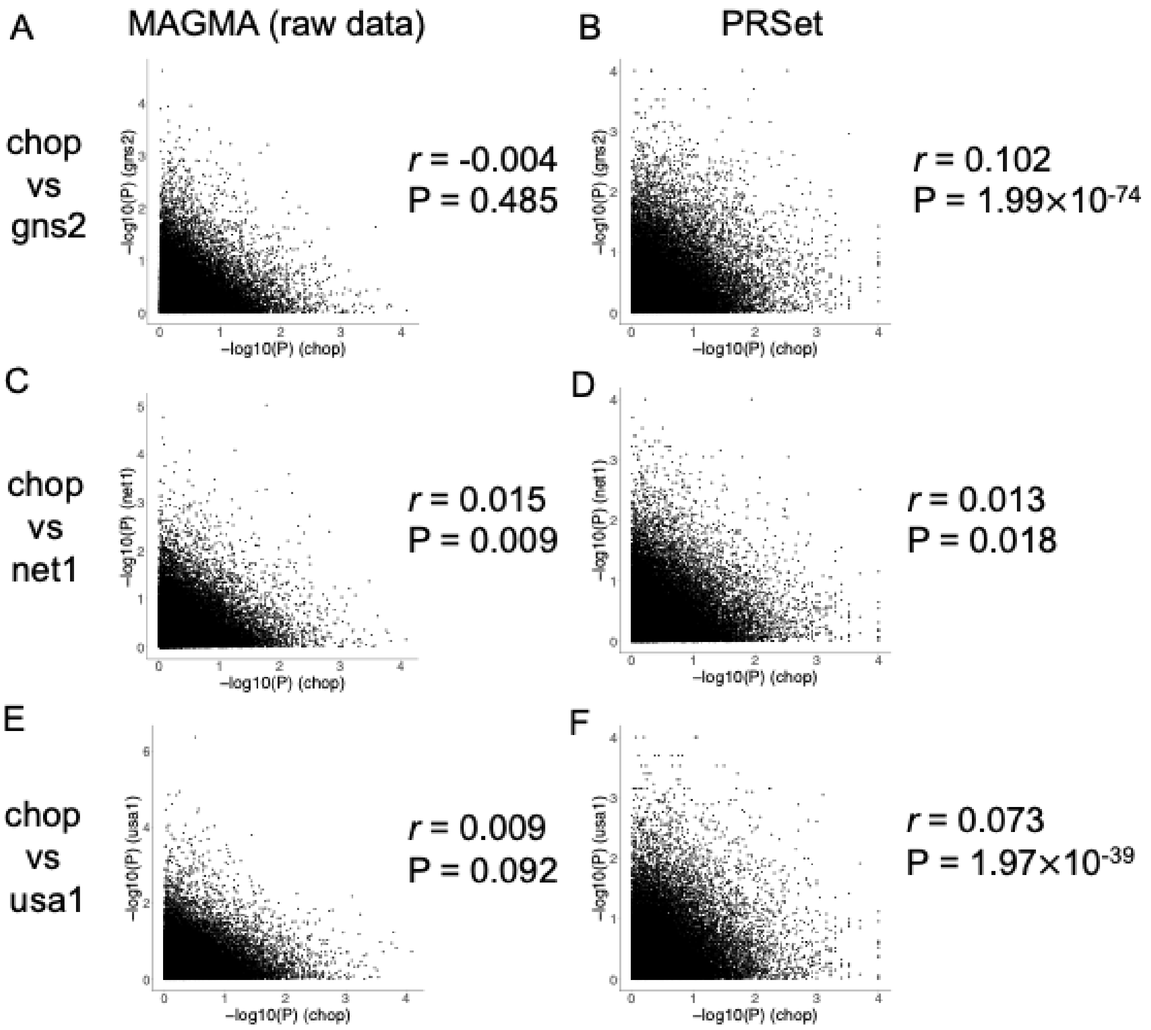


**Supplementary Figure 21. Neither MAGMA or PRSet has a high correlation of gene set p-value across discovery-replication cohort pairs.** Individual genetic data in these cohorts were used for gene set analysis by PRSet and MAGMA. Pearson correlation and its associated p-value are shown for each plot.

(A) Correlation between negative log10 p-value of the association between pathway and AN status in chop and negative log10 p-value in gns2 using MAGMA.
(B) Correlation between negative log10 p-value of the association between pathway and AN status in chop and negative log10 p-value in gns2 using PRSet.
(C) Correlation between negative log10 p-value of the association between pathway and AN status in chop and negative log10 p-value in net1 using MAGMA.
(D) Correlation between negative log10 p-value of the association between pathway and AN status in chop and negative log10 p-value in net1 using PRSet.
(E) Correlation between negative log10 p-value of the association between pathway and AN status in chop and negative log10 p-value in usa1 using MAGMA.
(F) Correlation between negative log10 p-value of the association between pathway and AN status in chop and negative log10 p-value in usa1 using PRSet.
Abbreviation: AN=anorexia nervosa.
